# Concurrent tDCS-fMRI: Impact of the current-induced magnetic fields on the measured BOLD signal

**DOI:** 10.64898/2026.06.04.26354901

**Authors:** Teresa Cunha, Miro Grundei, Fróði Gregersen, Till Nierhaus, Lars G. Hanson, Felix Blankenburg, Axel Thielscher

## Abstract

**Background:** Understanding how transcranial direct current stimulation (tDCS) affects brain activity critically benefits from the use of functional magnetic resonance imaging (fMRI) to measure the related BOLD (blood-oxygenation-level-dependent) signal changes. However, the small magnetic fields induced by the stimulation currents can cause artifacts in the fMRI images that can compromise findings from concurrent tDCS-fMRI studies.

**Objective:** To identify how the current-induced magnetic fields affect fMRI data and establish a quantitative framework for evaluating their impact on concurrent tDCS-fMRI measurements.

**Methods:** Magnetic fields induced by currents inside the head and electrode cables were calculated for a standard motor cortex montage. Their effects on echo-planar images (EPI) were simulated based on a framework derived from MR physics first principles and validated using phantom experiments. The framework was applied to artificially induce artifacts related to the tDCS current flow in current-free fMRI time series from 5 participants. These were compared to active runs from the same participants where tDCS intensity was varied in a block design.

**Results:** Currents in the electrode cables were the main contributors to the current flow-related artifacts in the EPI images, which occurred both locally by causing geometric distortions and remotely by affecting the dynamic update of the scanner demodulation frequency. The artificially induced fMRI activations corresponded well to those measured during real tDCS on the single-subject level for intensities of 2 mA and higher.

**Conclusion:** The current-induced magnetic fields can cause intensity changes comparable to typical BOLD responses. Their impact on the statistical results depends on the chosen experimental design (electrode locations, cable paths, imaging parameters, fMRI paradigm). The simulation framework provides a principled approach to evaluate the impact of these artifacts during the design and data analyses of concurrent tDCS-fMRI studies.

## 1. Introduction

Transcranial direct current stimulation (tDCS) is a well-established non-invasive brain stimulation (NIBS) technique, which applies weak direct currents (≤ 4 mA [1]) via scalp electrodes to modulate cortical excitability [2]. Functional magnetic resonance imaging (fMRI) plays an important role in understanding the physiological effects of tDCS at the brain-network level, and its use has grown substantially over the last decade [3–7]. fMRI captures changes in the blood-oxygenation-level-dependent (BOLD) signal, providing an indirect measure of neural activity associated with tDCS effects. Applying tDCS inside the MR scanner enables simultaneous assessment of both the immediate effects during stimulation and the aftereffects following stimulation.

Combined tDCS-fMRI studies pose specific requirements on the tDCS equipment to ensure participant safety and the quality of the fMRI data [6–9]. Because the BOLD signal is small, typically representing only a 1–5% change in MR signal intensity, the tDCS hardware must be optimized so as not to compromise the sensitivity of the fMRI measurements. For example, electrode materials should be selected to minimize susceptibility-related signal dropouts in nearby brain regions [4,10,11]. In addition, radio-frequency (RF) filters on the cables connecting the electrodes to the stimulator are necessary to prevent RF-induced noise artifacts in the fMRI data [12,13].

So far, it has been unclear whether the weak tDCS currents themselves can also affect the fMRI measurements to a relevant extent. Such effects might be particularly challenging, because changes of the tDCS intensity during fMRI acquisition, e.g. in a block design, could then cause systematic biases in the statistical results that are difficult to control for (see [14] for a related example for the combination of transcranial magnetic stimulation with fMRI). A concurrent tDCS-fMRI study in post-mortem subjects indeed revealed significant fMRI signal changes that were approximately half of the BOLD response during a finger tapping experiment in a healthy subject with identical block design and imaging sequence [8]. The study also simulated the tDCS current flow in the head and the resulting magnetic fields according to the Biot-Savart law. It reported a good spatial correspondence between the regions with the highest simulated current flow (and current-induced magnetic fields) and the strongest measured fMRI signal intensity changes, that both occurred mostly under the electrodes. It was concluded that the current-induced magnetic fields might have locally changed the main magnetic field of the scanner, causing the observed fMRI signal changes. However, the study was limited to qualitative comparisons, and it remained unclear whether the current-induced magnetic fields that are in the order of a few nanotesla (nT) can indeed exert a sufficiently strong impact. Further strong fMRI signal intensity changes were also measured in the ventricles that did not coincide with the simulated field peaks. Also, most effects were measured in superficial cerebrospinal fluid (CSF), potentially reducing the relevance of the findings for in-vivo tDCS-fMRI of the brain. In contrast, a later study estimated the potential BOLD confounds due to the magnetic fields induced by tDCS current flow in the head to be negligible [15]. This conclusion was derived from worst-case calculations of fMRI signal loss due to increased intravoxel dephasing, arising from intravoxel magnetic field gradients produced by the current-induced magnetic fields during stimulation.

Here, we developed a quantitative framework grounded in MR physics first principles that enables the explicit calculation of how magnetic fields generated by the tDCS currents influence the MRI signal. We stringently validated the framework by comparison with the MR signal measured in a series of phantom experiments. This allowed us to identify the main mechanisms through which current-induced magnetic fields cause artifacts in the fMRI time series. We then demonstrated that – when the tDCS current intensity is changed in a block design – the current-induced artifacts can explain a substantial part of the BOLD signal changes measured in humans. Specific experimental choices strongly influence the strength of the current-induced artifacts and their impact on the results of the statistical fMRI analyses. We therefore provide a simulation framework as a principled approach that enables researchers to evaluate the impact of the artifacts during the design and data analysis of concurrent tDCS-fMRI studies. The simulation scripts and an example dataset are publicly available at https://doi.org/10.17605/OSF.IO/GU6ER.

## 2. Methods

### 2.1. Participants

Five healthy volunteers (2 female) with no MRI contraindications were included in this part of the study after written informed consent. The study complied with the Helsinki Declaration on human experimentation and was approved by the Ethics Committee of the Capital Region of Denmark (De Videnskabsetiske Medicinske Komitéer, approval number 2206924).

### 2.2. tDCS-MR setup and data acquisition

Scanning was performed on a 3T MRI scanner (MAGNETOM Prisma, Siemens Healthcare, Erlangen, Germany) using a 64-channel head coil. tDCS was applied using a DC-STIMULATOR MR stimulator (neurocare Group GmbH, München, Germany) controlled by a custom signal generator that reads trigger pulses from the MR scanner to synchronize the current injection scheme with the MR pulse sequences. The stimulator was placed outside the scanner room and connected to the electrodes (or to a cable loop) via an RF filter built into the penetration panel. Inside the scanner bore, we used low-conductivity cables and electrodes made from silicon rubber that had the same material properties as those validated in our previous study [9]. Specifically, the cables were shown not to couple with the RF field, irrespective of the specific chosen cable length (see Fig 4 in [9]), ensuring safety and minimizing RF field distortions that affect SNR. Pilot experiments showed no changes in RF noise levels when the stimulator was connected and the electrodes and cables placed in the scanner.

The fMRI data of the human participants was acquired using the multi-band EPI sequence from the Center for Magnetic Resonance Research (Minneapolis, USA) [16], with the following parameters: repetition time T_R_^human^=1500 ms, echo time T_E_=37 ms, EPI echo spacing t_esp_=0.54 ms, flip-angle α=60°, 30 slices, 2D matrix size=110×110, 2 mm isotropic voxels, multiband factor 3, no RF spoiling and no in-plane acceleration. A gradient-echo field map (echo times T_E_=[4.92, 7.38] ms) was measured with the same field of view (FoV) and resolution as the EPI images for characterizing static B_0_ field inhomogeneities. A high resolution (0.9 mm isotropic) 3D ultra-short T_E_ structural scan (PETRA [17]) was additionally acquired to image the silicon rubber electrodes and/or cables. This allowed accurate reconstruction of the electrodes and cable paths in the latter analysis. Finally, standard T1- and T2-weighted anatomical scans (0.9 mm isotropic resolution) were acquired.

Additional data was acquired in a cylindrical phantom (diameter/height=110/120 mm) composed of an agarose gel (with added sodium chloride) surrounding three elongated water-filled compartments. These were doped with varying levels of copper sulfate, changing their relaxation rates relative to the background region of the phantom and providing contrast edges in the acquired MR data (Fig. 2A&F). The above settings for the EPI sequence were used, except that the repetition time was set to T_R_^phantom^=1000 ms, no multiband acceleration was applied, and the number of slices was reduced to 11. One aim of the phantom tests was to assess the effect of the current flow on the updates of the demodulation frequency that the scanner performs to compensate for frequency drifts over time [18]. We therefore disabled the online updates of the demodulation frequency in some of the acquisitions (in which case the MR scanner applies an automatic offline correction to the DICOM images instead), and reconstructed the EPI images from raw k-space data using our own software to get time series not affected by the frequency updates. Multiband acceleration was disabled to enable reconstruction using our software.

### 2.3. Stimulation paradigm

Two setups were tested in the human participants (Fig. 1A): Real tDCS was applied using a montage with circular electrodes (diameter=35 mm) placed over the left motor cortex (M1) and contralateral supraorbital area (SO) [2]. The electrodes were attached to the skin using Ten20 conductive paste (Weaver and Company, Aurora, CO, USA), following recommended procedures [1,6] to ensure good contact. The electrode cables were led in the superior direction through an opening in the coil, similar to prior tDCS-fMRI studies [9,12,19]. Periods of positive and negative currents were applied in a block design (Fig. 1B) with ramp and plateau durations of 3 and 18 seconds, respectively. The succession of baseline and stimulation periods was optimized to minimize sensitivity to low-frequency signal fluctuations. Separate fMRI acquisitions were performed without stimulation (0 mA) and for three current intensities (1, 2 & 3.5 mA). Additional acquisitions were performed for currents flowing in a cable loop around the head, using two current intensities (1 & 2 mA) and without current flow (0 mA). This enabled testing the effects of the current-induced magnetic field on fMRI data without confounding physiological stimulation effects [20]. The relevant component of the current-induced magnetic field that is parallel to the main scanner field (z-direction per convention) is denoted by ΔB_zc_ in the following.

**Figure 1:**
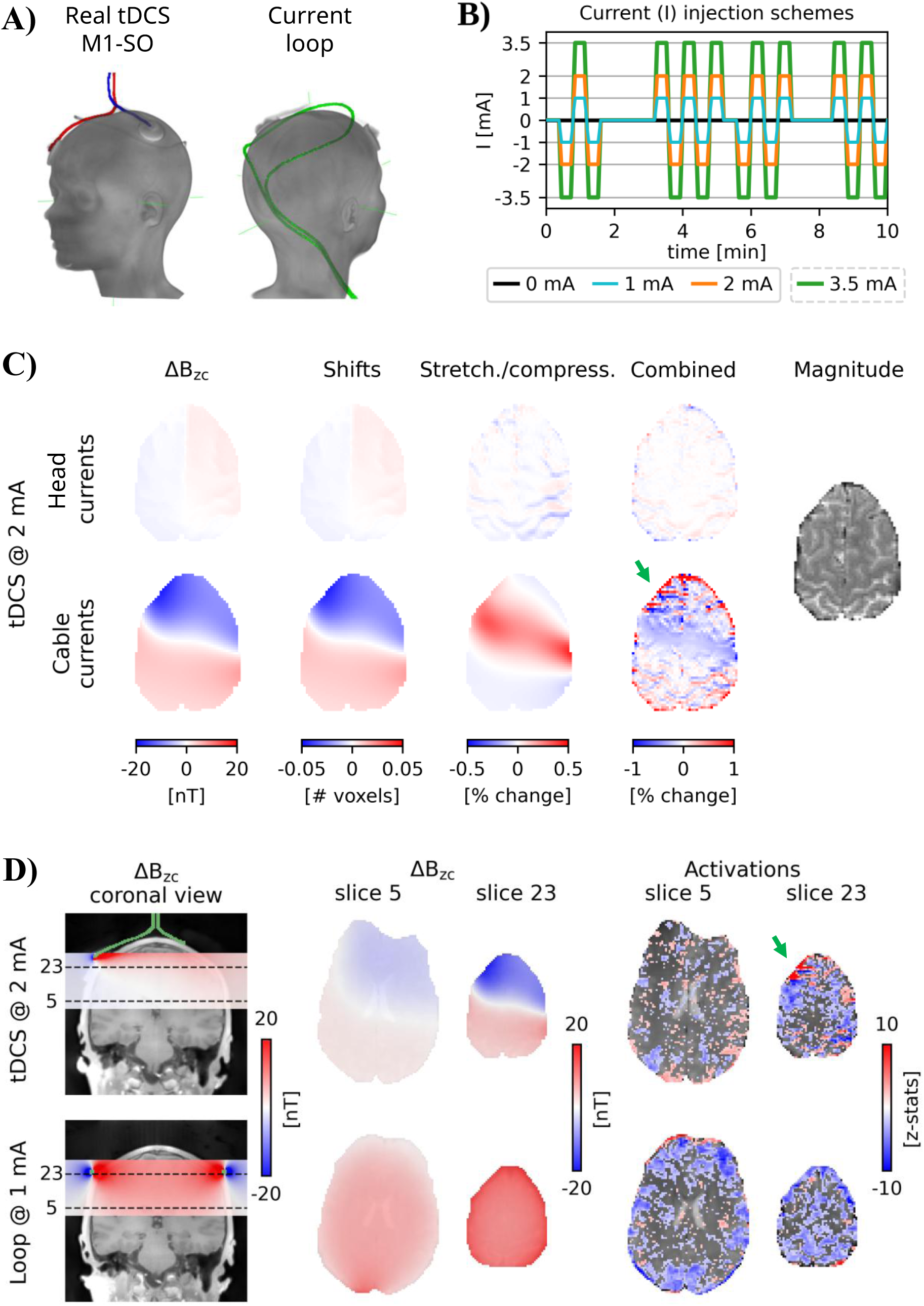
Effects of the current-induced magnetic fields (ΔB_­_) on echo planar images (EPI). **A)** PETRA scans showing the electrode placement and lead paths in a real tDCS experiment and the lead path in a current loop experiment. The colored leads were manually tracked in the scans. **B)** Current injection schemes. Different current intensities were used in the tDCS and loop experiments to induce magnetic fields of comparable intensity. **C)** Predicted geometric EPI distortions for the real tDCS experiment in a slice close to the electrodes (slice 23). First row: Effects due to the magnetic fields induced by the currents in the head. Second row: Effects due to the current flow in the electrode leads. From left to right: ΔB_­_estimates based on the Biot-Savart law and current flow calculations inside the head or the lead paths; maps of the ΔB_­_–induced shifts; maps of the intensity changes from the concomitant stretching/compression; percentage intensity change from those two effects combined; corresponding magnitude image. **D)** Current-induced effects in resting-state fMRI performed during real current injection (top) or current flow in a cable loop (bottom). From left to right: Simulated magnetic fields of the currents flowing in the electrode leads (highlighted in green) or cable loop in a coronal slice; simulated magnetic fields in two transversal slices; activation maps from first-level fMRI analyses of the current injection schemes shown in subfigure **B** (z-statistics thresholded at 1.65).

In the measurements with a phantom, a cable loop was placed above it and the current intensities changed in a block design (Figs. 2A&C). Additional acquisitions were performed without current flow (0 mA).

### 2.4. Preprocessing and analysis of the fMRI data

The human fMRI data was corrected for movement using the rigid motion correction method implemented in FSL MCFLIRT [21], thereby using the first volume of the initial fMRI acquisition as a reference to spatially co-register all acquisitions. First-level fMRI analyses were conducted in FSL FEAT [22]. Temporal high-pass filtering with a 50-second cutoff period was applied to the motion-corrected series and to the regressors in the design matrix. These consisted of the current injection scheme (split into two orthogonal regressors of interest, which included either the negative or the positive blocks) and the motion parameters as co-regressors of no interest. The splitting of the current injection scheme into positive and negative currents accounts for the fact that symmetric current-induced magnetic fields do not necessarily result in perfectly symmetric intensity changes. Prewhitening was not applied unless stated otherwise. The analysis was performed on either non-smoothed data, or after smoothing with 3 or 6 mm FWHM (full-width-at-half-maximum) kernels. The 6 mm smoothed data was additionally analyzed using a version of the current injection scheme that was convolved with the HRF, replicating a standard BOLD-fMRI analysis.

For the phantom fMRI data, temporal high-pass filtering with a 100-second cutoff period was used. Neither motion correction nor spatial smoothing was applied. For the first-level fMRI analyses, the current injection scheme was split into two orthogonal regressors of interest, which included either the negative or the positive blocks.

### 2.5. Simulation of the current-induced magnetic fields and their impact on the EPI images

For simulating the ΔB_zc_ fields caused by the current flow in the cables, we manually tracked the cable paths in the acquired PETRA images that were co-registered to the first volume of the motion-corrected EPI series using FSL FLIRT. The ΔB_zc_ fields were then numerically calculated using the Biot-Savart law. The accuracy of the simulated cable-induced ΔB_zc_ fields was cross-validated with MR current density imaging in phantom measurements (Supporting Material S3).

In one participant, we additionally simulated the ΔB_zc_ field due to the current flow induced by the M1-SO tDCS montage inside the head using SimNIBS 4.0.0 [23]. For that, we created a head model from the structural T1- and T2- images using the *charm* pipeline. The T1 image was rigidly co-registered to the EPI images, and the transformation applied to the head model. After simulation of the current flow using the finite element method (FEM) of SimNIBS, the resulting ΔB_zc_ fields were again calculated using the Biot-Savart law.

We then calculated the geometric distortions caused by the ΔB_zc_ fields in the EPI images, in dependence on the chosen echo spacing. The corresponding theory and its experimental validation are described in Supporting Material S1. In short, EPI imaging is known to be prone to local geometric distortions consisting of voxel shifts in the phase-encoding direction and intensity changes from the associated stretching/compression due to inhomogeneities of the main B_0_ field. These effects are typically seen in regions of strong magnetic susceptibility gradients (e.g., close to air-tissue interfaces) [24,25]. The ΔB_zc_ fields caused by the currents add to the B_0_ inhomogeneities and thus affect the EPI images via the same mechanisms, but to a smaller extent given their strength in the nanotesla range.

We also calculated the effects of the ΔB_zc_ fields on the automatic updates of the demodulation frequency that the scanner performs to compensate for frequency drifts over time, e.g., due to heating of the gradient and shim coils [18]. The related theory and its validation are described in Supporting Material S2. Put briefly, the scanner measures the average response frequency of the hydrogen nuclei in the FoV and uses this to adjust the demodulation frequency during image acquisition. The response frequency is linearly related to the main B_0_ field via the Larmor equation, and B_0_ changes due the current-induced ΔB_zc_ fields will thus lead to changes of the demodulation frequency that show up as global image shifts in the phase-encoding direction in case the ΔB_zc_ fields are sufficiently strong.

We applied the ΔB_zc_-induced geometric distortions and effects on the demodulation frequency updates to the EPI time series acquired without stimulation, thereby replicating the temporal changes of the stimulation currents in the block designs (Figs. 1B&2C). For that, we brought the simulated ΔB_zc_ into the same “distorted space” as the EPI images using the acquired GRE B_0_-field map as a forward distortion map. The expected local geometric distortions were applied according to the theory in Supporting Material S1. The effects of the demodulation frequency updates were simulated via global image shifts in the phase-encoding direction, as described in Supporting Material S2. We then compared the EPI time series with artificially induced artifacts with the fMRI data acquired with real current injection. For the human fMRI data, this step was applied before motion correction.

Finally, we also estimated the ΔB_zc_ effects on intravoxel dephasing (Supporting Material S4), confirming that the signal loss due to this mechanism is negligible [15]. We further evaluated the impact of the ΔB_zc_ fields on the motion correction performed during fMRI analysis (Supporting Material S5). While we could find an influence on EPI data recorded from the phantom, the effects on the human EPI data were very small compared to the impact of real movements. We thus used the standard motion correction of FSL (MCFLIRT) for the main results.

### 2.6. Accuracy assessment of the artifact simulations

We quantitatively evaluated the accuracy of the simulated artifacts by comparing the statistical activation maps (i.e., z-statistics) obtained for the fMRI data with simulated artifacts added and data recorded for real current flow. We used linear fits to the voxel-wise z-values with the results for real current flow and simulated artifacts as independent and dependent variables, respectively. Since both results contained measurement noise of comparable variance, we used the errors-in-variables (Deming) regression with variance ratio equal to one [26,27] instead of ordinary least squares regression. The coefficient of determination (R^2^) was computed according to the generalized R^2^ proposed in [27]. This is a conservative choice, as it represents the minimum percentage improvement of the Deming regression line over the horizontal and vertical lines that pass through the centroid of the data.

We performed permutation tests to quantify the significance of the linear fits on the single-subject level. Our hypothesis was that adding the simulated artifacts to the 0 mA data substantially improves the R^2^ of the fits to the data with current flow, compared to the original 0 mA without artifacts. To test this formally, we generated a null distribution *H_0_* of R^2^ values that would be obtained if there was no systematic difference between the activation maps determined for 0 mA data with and without simulated artifacts. This was achieved by randomly exchanging voxels between the two maps and calculating new linear fits to the maps for real current flow. This procedure was repeated 100,000 times. One-sided p-values were then computed as the fraction of random samples that were larger than the R^2^ of the fit between the results determined for simulated and real current-induced artifacts. Significance was assessed at p=10^-5^ (uncorrected).

## 3. Results

In the following, we first demonstrate by which mechanisms the current-induced magnetic fields cause artifacts in fMRI data (3.1). We show that different mechanisms contribute to artifacts that occur at positions where the current-induced magnetic fields are strong and at remote locations, respectively. We then quantitatively simulate the artifacts in EPI time series that were recorded without current flow and confirm that they essentially match the true effects measured both in phantom and human data (3.2).

### 3.1. Characterizing the current-induced artifacts in tDCS-fMRI

As first step, we calculate the expected effects of the current-induced magnetic fields ΔB_zc_ caused by the M1-SO tDCS montage in one of the participants in an EPI slice close to the electrodes for currents of 2 mA (Fig. 1C). The simulated ΔB_zc_ from the current flow in the electrode cables are much stronger (reaching 20 nT close to the electrodes) than those induced by the diffuse current flow inside the head (up to 2.5 nT). This translates into stronger ΔB_zc_-induced voxel shifts and intensity changes from the associated local stretching or compression. Thus, we only consider the ΔB_zc_ of the cables in the following. While stretching/compression directly changes the image intensity, the strongest intensity changes result from sub-voxel shifting, since it leads to changes in partial volume effects at the boundaries between tissues with high intensity contrast (e.g., around the sulci). Local intensity changes of up to 1% are expected for a current intensity of 2 mA in the electrode cables, which are comparable to typical BOLD responses [28]. In contrast, the predicted changes due to the current flow inside the head are about an order of magnitude lower.

Visually comparing the activation maps from first-level fMRI analyses of resting-state data acquired during real tDCS (first row of Fig. 1D; current injection scheme in Fig. 1B) shows that the measured activations in the slice close to the electrodes have a similar spatial pattern as the predicted intensity changes due to the geometric distortions caused by the cable fields (data highlighted by the green arrows), suggesting that the activations were influenced by artifacts. Strong activations were also observed for currents of 1 mA flowing in a loop around the head (Fig. 1D second row), which induced magnetic fields of comparable intensity to those in tDCS. This further corroborates the hypothesis that the spatially concentrated current flow in cables results in strong magnetic fields that can result in artifacts in fMRI data. Importantly, the effects observed in slice 5 distant from the cable loop suggest that different artifacts occurred in this case in addition to local geometric distortions in the regions of the strongest magnetic fields.

In order to understand the causes of these remote artifacts, we tested the impact of ΔB_zc_ on the automatic updates of the demodulation frequency performed by the MR scanner in a phantom (Fig. 2). Time series of the frequency offsets in two slices – close and remote to the cables – are shown in Fig. 2E, for acquisitions with and without frequency updating. We reconstructed the data without frequency updating from raw k-space data using our own software, as the DICOM files provided by the scanner are always automatically corrected for frequency drifts. The unprocessed measurements (first rows) reveal the expected frequency drift due to technical causes such as gradient heating over the course of the EPI time series when the frequency updates are not applied. After detrending (second rows), a contribution of ΔB_zc_ to the frequency offsets is clearly visible. Importantly, comparing the left and right detrended plots reveals how ΔB_zc_ affects the corrections made by the scanner: The “strong” ΔB_zc_-induced frequency offsets in locations close to the cable (e.g., slice 9) got reduced, while the weak offsets far from the cable (e.g., slice 1) got inverted and amplified. The impact of the undesired effects of ΔB_zc_ on a first-level fMRI analysis of the current injection scheme are shown in Fig. 3F. In general, it causes global image shifts in the phase-encoding (A-P) direction, visible as artifactual activations at object edges. Without frequency updates, the artifacts are mostly restricted to slice 9 close to the cable loop, caused by local geometric distortions. When frequency updates are applied, they partially compensate for the local artifacts in slice 9 (green arrows) but introduce additional remote artifacts in slice 1 (red arrows) by shifting the entire phantom in the A-P direction.

**Figure 2:**
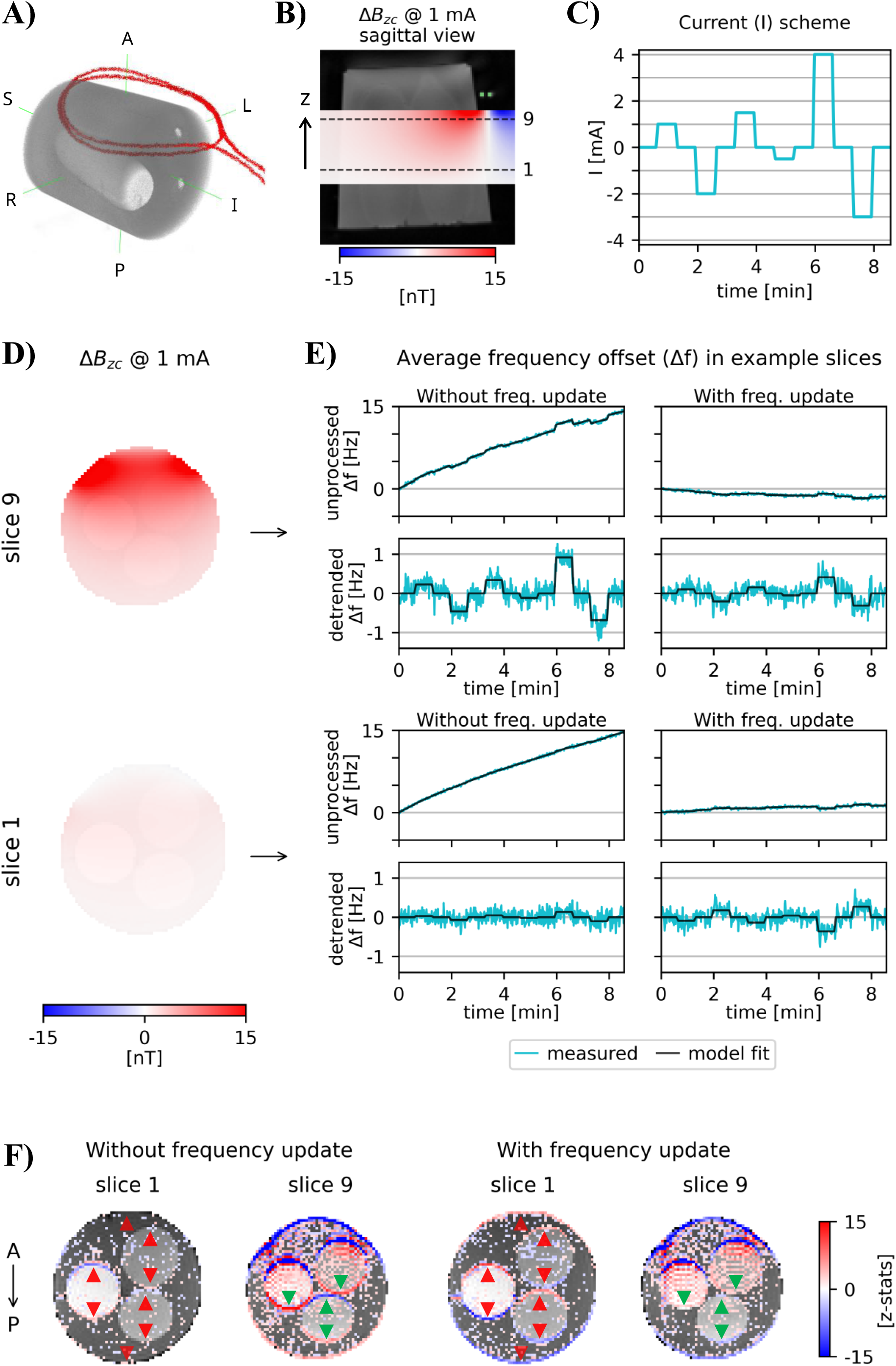
Sensitivity of the demodulation frequency updates to the current-induced magnetic fields (ΔB_zc_). **A)** Visualization of the in-house-built phantom and cable loop. **B)** Sagittal view of ΔB_zc_ calculated for 1 mA and shown within the field-of-view (FoV) of the EPI data, overlaid on a T1-weighted image of the phantom. The magnetic field increases along the slice direction (z) with decreasing distance to the conductor on top. The dashed lines indicate the 1^st^ and 9^th^ EPI slices. **C)** Current waveform. **D)** ΔB_zc_ simulations in the two EPI slices, overlaid on the first EPI image of the time series. Note that ΔB_zc_ was substantially weaker in slice 1, which was further away from the cable. **E)** Time series of the frequency offset in the two slices for acquisitions without (left) and with (right) online updating of the scanner’s demodulation frequency. For each slice, both unprocessed (top) and detrended (bottom) frequency offset measurements are shown. The unprocessed measurements reveal the expected frequency drift over the course of the EPI time series when the online updates were disabled. The detrended series show a clear contribution of ΔB_zc_ to the frequency offset. Comparing the left and right plots shows how ΔB_zc_ affects the frequency updates applied by the scanner: The ΔB_zc_-induced frequency offsets in slice 9 (close to the cables) get reduced, while the offsets in slice 1 (far from the cable) get inverted and amplified. **F)** Activation maps from a first-level fMRI analysis of the current injection scheme shown in subfigure **C** (z-statistics thresholded at 1.65). Results are shown for acquisitions without (left) and with (right) frequency update. The frequency update caused global image shifts, which resulted in signal changes at edges of the phantom and inner compartments due to changes in partial volume effects. These added to the local signal changes due to the geometric distortions, partially compensating them in slice 9 (green arrows) and introducing new ones in slice 1 (red arrows).

**Figure 3:**
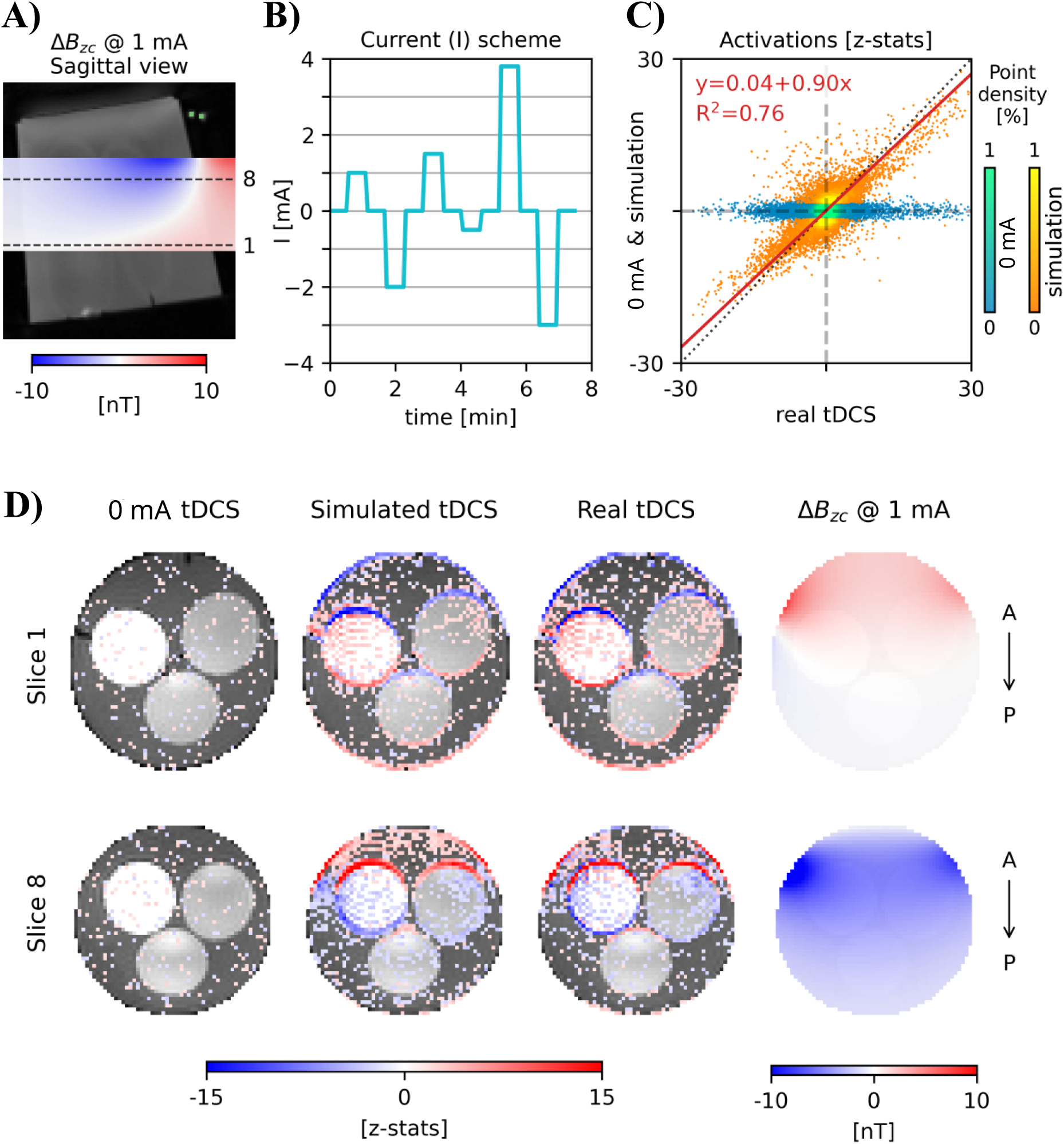
Simulation of the artifacts caused by the current-induced magnetic fields (ΔB_zc_) in a phantom. **A)** Sagittal view of ΔB_zc_ calculated for 1 mA and shown within the FoV of the EPI data. The cable configuration was similar to the one in Figure 2A. The dashed lines indicate the 1^st^ and 8^th^ EPI slices. The green dots show the positions of parts of the cable. **B)** Current waveform. **C)** Scatter plots of the activations (z-statistics) from first-level analyses of sham and simulated tDCS, against activations determined in the same way for real tDCS. The blue point cloud for sham vs real tDCS is horizontally elongated. Adding simulated tDCS artifacts to the 0 mA EPI time series makes them similar to the data acquired during real tDCS. Correspondingly, the linear fit to the orange point cloud shows a high correlation coefficient and a slope near 1. The z-statistics of the simulated tDCS were thresholded at 1.65 before computing the linear fit between real and simulated activations (red line). **D)** Activation maps for the two slices (z-statistics thresholded at 1.65) for 0 mA, simulated and real tDCS, and simulated ΔB_zc_ overlaid on the first image of the EPI time series.

### 3.2. Comparing simulated current-induced artifacts with recorded tDCS-fMRI data

As first step, we calculated the expected artifacts caused by geometric distortions and effects on the frequency updates due to current flow in a cable loop around a phantom (current injection scheme in Fig. 3B). Artificially inducing them in an EPI time series recorded without currents resulted in artifactual activations that were very similar to the results recorded for real current flow (Fig. 3D). The remaining differences seem to mainly be due to the measurement noise inherent in both the sham and real EPI time series. The accuracy of the simulations is confirmed by the strong linear relationship between the activations determined for simulated and real tDCS (orange point cloud in Fig. 3C) that shows an intercept close to zero, a slope approaching one and a high R^2^ value (significant at p<10^-5^ uncorrected). As statistical results are usually interpreted after applying a threshold, we used a lenient voxel-level threshold of z>1.65 for the simulated tDCS data (corresponding to one-sided p<0.05). However, we confirmed that this choice is not critical; the strength of the linear relationship increases for higher thresholds. Please also note that we did not apply multiple comparison correction to the data as our aim was to evaluate the similarity of the spatial distributions of voxel-wise z-values. However, the z-values at the contrast edges are sufficiently high to survive conservative Bonferroni correction (e.g. z=4.75 corresponds to p=10^-6^). The blue point cloud shows the 0 mA data before inducing the artifacts, plotted against real tDCS, confirming the absence of a systematic relationship.

We then assessed the relationship between data measured during real current flow and simulated artifacts in five participants, both for current flowing in the cables of a M1-SO tDCS montage and in a loop around the head. The results in the following are focused on the upper half of the acquisition field-of-view (FoV) in which the currents in the cables of the real tDCS induced the strongest ΔB_zc_ fields and the artifacts were mostly due to local geometric distortions (slice 16-28 in Fig. 4A). Figs. 4B&C show the results for an example subject (subject 1) for the M1-SO tDCS montage with an intensity of 3.5 mA and for the current loop with 2 mA. The results were analyzed without HRF convolution and for a spatial smoothing of 3 mm FWHM. In both cases, a good correspondence between the fMRI measurements during real current flow and simulated ΔB_zc_ artifacts added to 0 mA data can be visually observed in the activation maps from the first-level analyses and are confirmed by the linear fitting results (significant at p<10^-5^ uncorrected). As expected, the blue point clouds that show 0 mA plotted against real activations have a wider spread in the vertical direction than observed in the phantom (Fig. 3C). This is caused by higher baseline noise levels dominated by physiological noise in the human measurements, and can reduce the correlation between the EPI time series obtained during real stimulation and simulated artifacts in the human results. The correlation nevertheless stays high.

**Figure 4:**
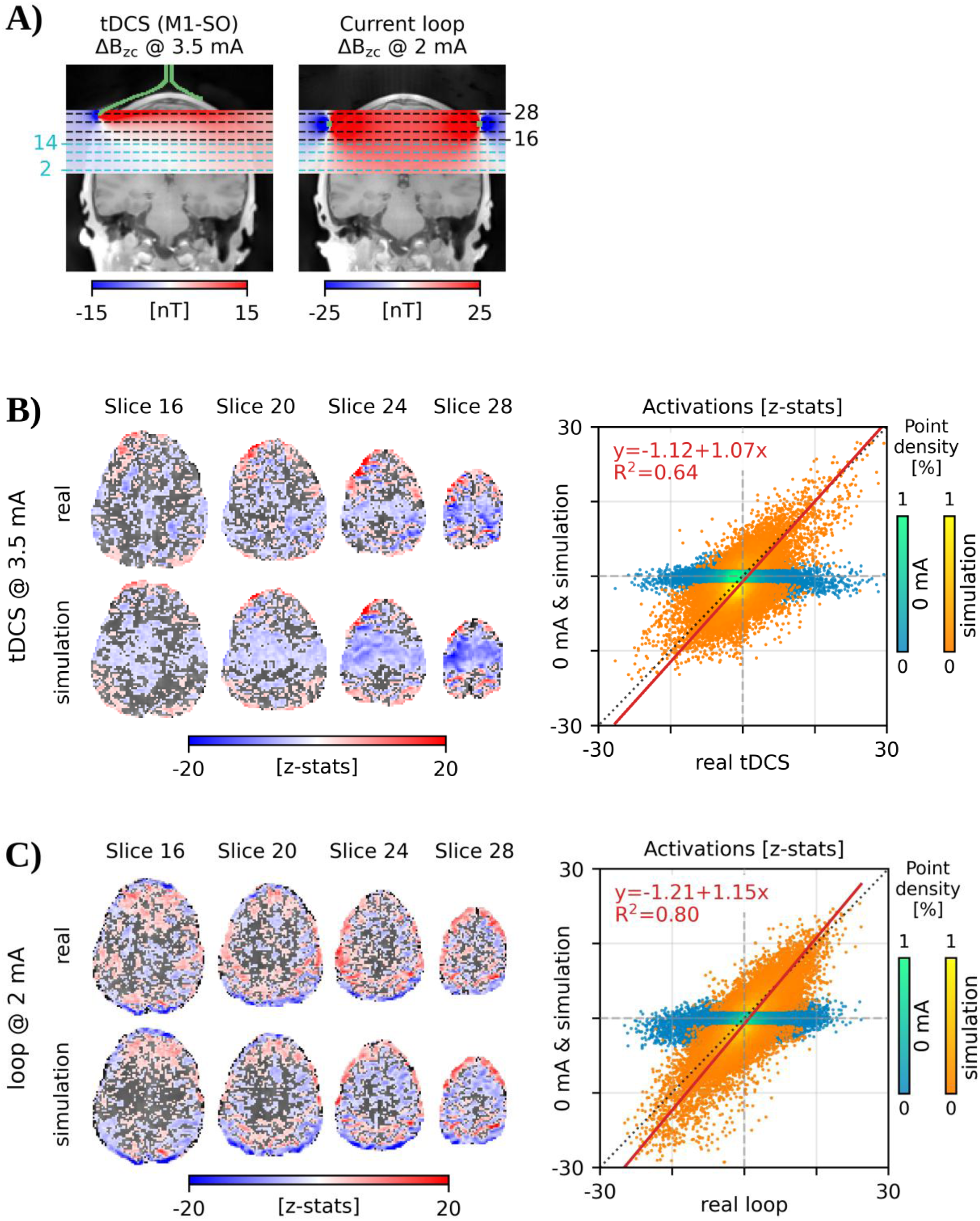
Simulation of the artifacts caused by the current-induced magnetic fields (ΔB_zc_) in a human brain, for both real tDCS (M1-SO montage) and current loop. **A)** Simulated ΔB_zc_ for 3.5 mA and 2 mA currents flowing in the electrode leads and cable loop, respectively. The simulations are shown in the FoV of the EPI data, overlaid on a coronal slice of a PETRA image. The dashed black lines depict the upper EPI slices chosen for further illustration below, while in cyan are the lower slices shown in Supplementary Figure S7. The green lines or dots depict the cable paths, which were similar to those in Figure 1A. Note that the magnetic fields scale linearly with the applied currents. **B&C)** Left: Activation maps (z-statistics thresholded at 1.65) for the real and simulated tDCS/loop in the four example slices for a current intensity of 3.5/2 mA, overlaid on the first image of the EPI series. Right: Scatter plot of the activations obtained for 0 mA and simulated tDCS/loop against real tDCS/loop, and linear fit between the simulated and real activations (red line) in the *upper* half of the FoV. The z-statistics of the simulated tDCS/loop were thresholded at 1.65 before computing the linear fit.

The results for all five participants and tested current intensities are summarized in Fig. 5. Shown are the slopes and R^2^ values of the linear fits to the voxel-level z-statistics determined for data obtained with real current flow and simulated artifacts. For the M1-SO tDCS montage (Fig. 5A), consistent relationships with R^2^ values close to or exceeding 0.5 are seen in all subjects for current intensities of 2 mA and 3.5 mA, irrespective of the chosen smoothing levels or thresholds (indicated on the lower and upper x-axes, respectively). They approached the maximally expected R^2^ values given the measurement noise (indicated by the horizontal dotted gray lines), which we estimated from additional measurements in 3 subjects with identical stimulation (Supporting Material S6). Replicating a standard fMRI analysis by using an HRF-convolved version of the current injection scheme as regressor on average reduced the strength of the correlations but did not abolish them. Repeating the analyses for pre-whitened data yielded very similar results (Supporting Fig. S12). The distributions of z-statistics had consistently longer tails than those observed for 0 mA, with a substantial number of voxels exhibiting z-values above 10 (Supporting Fig. S13). These corresponded to percent signal changes comparable to typical BOLD responses in task fMRI (Supporting Fig. S14). For a current intensity of 1 mA, consistent relationships were still observed in some participants.

**Figure 5:**
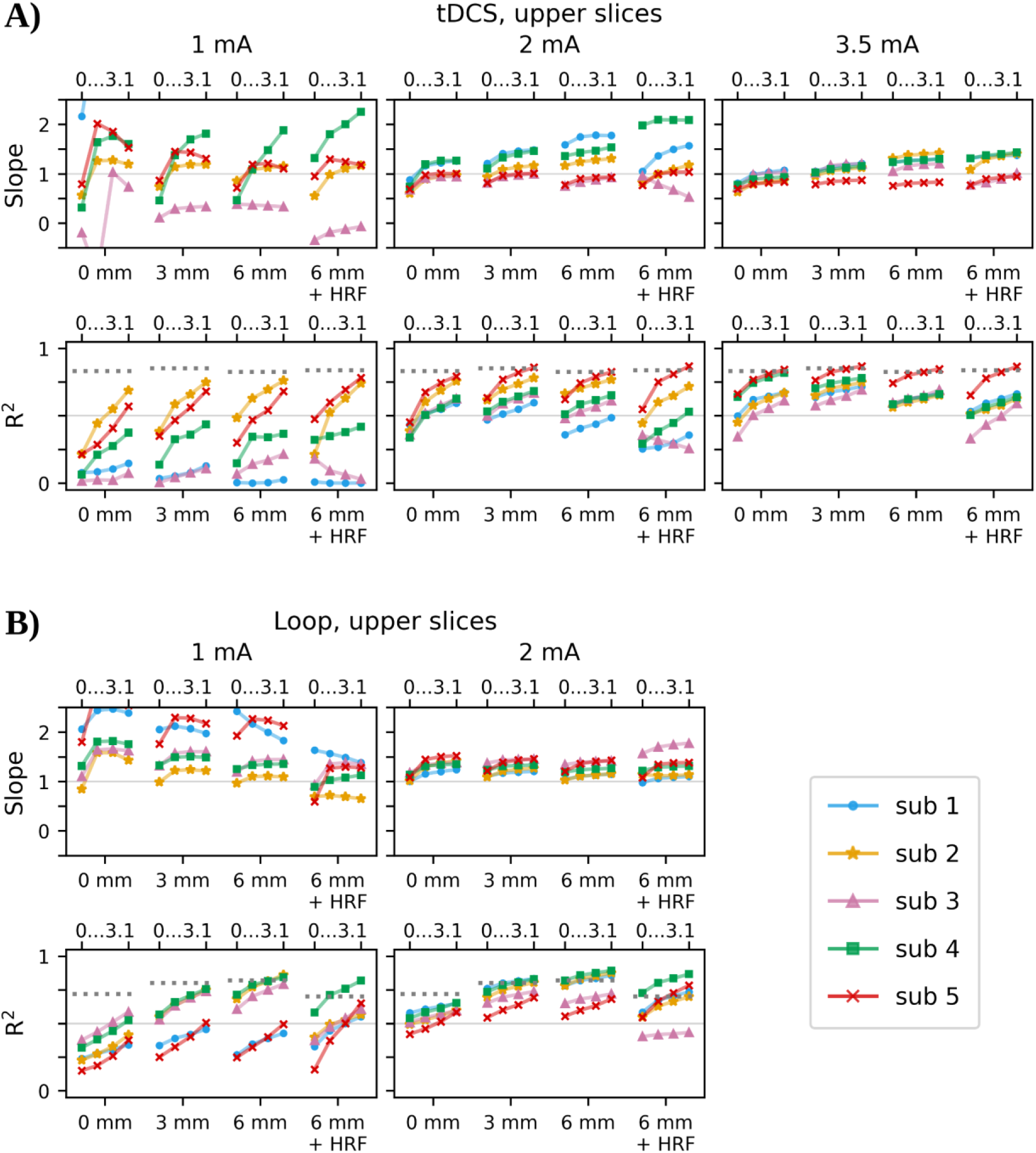
Summary results of the artifact simulation for all subjects and tested current intensities. Shown are the slopes and coefficients of determination (R^2^) of the linear fits to the z-statistics calculated for real and simulated current-induced artifacts for the EPI data in the *upper* half of the FoV. Spatial smoothing levels of the EPI time series were varied between 0, 3 and 6 mm FWHM, as indicated on the horizontal axes. The time series smoothed with 6 mm were additionally analyzed using a version of the current injection scheme that was convolved with the Haemodynamic Response Function (HRF), replicating a standard approach for BOLD fMRI analyses. The linear fits were evaluated for different thresholding levels (0, 1.65, 2.3 and 3.1; corresponding to one-sided p-values of 1.0, 0.05, 0.01 and 0.001) of the simulated data, indicated on the upper horizontal axes. The approximate upper bounds for R^2^ due to measurement noise are shown as horizontal dotted lines. **A)** tDCS results for 1, 2 and 3.5 mA. **B)** Loop results for 1 and 2 mA.

The results for the current loop (Fig. 5B) resemble those obtained for real tDCS, with consistent relationships observed for 2 mA on the single-subject level that are close to the maximally expected R^2^ values, and weaker relationships when using an HRF-convolved version of the current injection scheme as regressor or when reducing current intensity to 1 mA. These results confirm the correctness of our simulation framework and again demonstrate that the spatially concentrated current flow in cables can induce clearly measurable effects in EPI time series at current intensities used for tDCS.

We applied permutation testing to the R^2^ values obtained for 6 mm smoothing (with and without HRF convolution) and a threshold of z>1.65 for both real tDCS and the current loop. They were significant in 46 of 50 cases (tested at p=10^-5^, corresponding to p=0.01 Bonferroni-corrected for 50 comparisons). Exceptions are subject 1 for tDCS at 1 mA (with and without HRF convolution), subject 3 for tDCS at 1 mA (HRF convolution), and subject 4 for tDCS at 3.5 mA (without HRF convolution).

In the lower half of the FoV (slice 2-14 in Fig. 4A), the artifacts are mostly caused by effects of the frequency updates. The strength of these effects depends on the spatial average of the ΔB_zc_ field in the overall FoV. As positive and negative ΔB_zc_ fields mostly average out for the M1-SO tDCS montage (Fig. 1D – first row), this results in weak artifacts in the lower slices. For a current intensity of 3.5 mA, R^2^ values around 0.5 are achieved in 3 of the 5 participants (Supporting Fig. S11A). For the current loop around the head, the ΔB_zc_ fields in the FoV are consistently positive (Fig. 1D – second row). This results in clear remote artifacts with the R^2^ values exceeding 0.5 in all participants for a current flow of 2 mA. Consistent relationships still occurred in some participants for 1 mA (Supporting Fig. S11B).

Auxiliary group-level analyses revealed that the current-induced magnetic fields resulted in spatially systematic artifacts that did not average out across subjects (Supporting Material S8). Substantial linear relationships between real and simulated tDCS/loop were observed even at 1 mA (Supporting Fig. 15). While restricted to fixed-level analyses, the auxiliary results confirm that the artefacts can be expected to impact standard group-level fMRI analyses.

## 4. Discussion

We demonstrated that when applying tDCS during fMRI, the current-induced magnetic fields can change the measured fMRI signal to a similar extent as physiological BOLD activations. The magnetic field due to the current flow in the electrode cables causes local geometric distortions in nearby brain regions, resulting in intensity modulations due to stretching/compression and sub-voxel shifts at tissue boundaries. In addition, it can affect the updates of the demodulation frequency performed by the scanner, causing global image shifts and related artifacts at tissue boundaries in remote regions. In contrast, the current-induced magnetic field did not lead to signal changes due to intravoxel dephasing, in line with earlier findings [15]. The magnetic field due to the current flow inside the head was generally weaker and not the main cause of the artifacts.

We derived a quantitative framework based on MR physics first principles to simulate the current-induced magnetic fields in EPI time series and validated it in phantom measurements. We then confirmed that the current-induced magnetic fields contributed to the fMRI signal changes measured in five participants. To do so, we simulated the expected artifacts in the fMRI time series when changing the stimulation intensity in a block design for a M1-SO tDCS montage. The artificially induced fMRI activations corresponded well to those measured during real current flow on the single-subject level for intensities of 2 mA and higher. Their magnitude was in the range of true physiological BOLD activations. For an intensity of 1 mA, consistent relationships were still observed in some participants, while technical and physiological noise dominated over the artifact-related signal changes in the other cases. Repeating this approach for currents flowing in a cable loop around the head served as additional validation of the quantitative simulations for in-vivo measurements. Our study confirms the occurrence of artifactual fMRI signal changes that have been previously reported for tDCS in two post-mortem subjects [8]. It demonstrates that these artifacts can also occur in-vivo and identifies their root causes.

Our findings provide important insights that enable minimizing the artifacts and their impact on statistical fMRI analyses. The physical setup should be optimized to make the cable paths as parallel as possible to the main B_0_ field of the scanner to reduce the impact of their current-induced magnetic fields [9]. In contrast, the M1-SO tDCS montage tested here followed recommendations that aim to ensure the safety of commercial MR-compatible tDCS cables by minimizing their path lengths close to the subject and leading them out through a rear port of the head coil. However, this causes strong ΔB_zc_ fields. Using low-conductivity silicon rubber cables [9] that do not couple to the RF excitation field of the scanner allows keeping them better aligned to B_0_ by leading them out towards the front. However, depending on the electrode location, substantial bending of the cables may still be unavoidable.

The impact of the artifacts on the results of statistical fMRI analyses will depend on the chosen experimental paradigm. Here, we changed current intensities in a block design [4,29–34] and focused on single-subject level analyses to unambiguously identify the artifacts and validate the simulation framework. However, it seems likely that also standard group-level analyses will be affected when a similar design is used. Less impact is expected when, e.g., constant currents are applied throughout the fMRI acquisition [10,35], even though it seems still relevant to evaluate whether the artifacts might influence group-level comparisons between sham and active runs in this case. Transcranial alternating current stimulation (tACS) was not tested here, because its impact on the EPI signal is expected to be negligible as the current-induced magnetic fields periodically change their sign [8]. Nevertheless, our results suggest that the EPI repetition time should not be chosen as a multiple of the tACS current period to avoid that the same EPI slice (or groups of slices for SMS) is always acquired with the same tACS phase.

Evaluating the impact of the artifacts for different experimental designs and choices (e.g., cable paths, stimulation intensity, fMRI paradigm, imaging parameters, spatial and temporal filter settings) was beyond the scope of this study. Instead, we focused on deriving and validating a quantitative framework to simulate the artifacts. We suggest that it can guide the planning of optimized tDCS-fMRI experiments based on testing the impact of simulated artifacts on available in-house or openly accessible fMRI datasets. In addition, we suggest to use it for reaching more robust analyses results for recorded tDCS-fMRI data. Specifically, statistically comparing the fMRI activations obtained from data with real stimulation with sham data that contains artificially added artifacts enables evaluation of the robustness of the reported fMRI activations. This requires that the cable paths can be reconstructed on the individual level, which we did here by imaging the rubber cables using PETRA scans. Alternatively, the current-induced magnetic fields can be measured directly by means of MR current density imaging (MRCDI) [15,36–38] (Supporting Material S3 gives methodological details to obtain unambiguous MRCDI measurements). Our framework is generic and can be applied to EPI data from other vendors or acquired with different EPI sequences. While we consider it unlikely, the implementation details of the frequency updates applied by the scanner might change the impact of the current-induced magnetic fields on those updates. It would thus be useful to technically validate the predicted global image shifts in these cases, e.g., by measurements with a cable loop placed on a phantom (Supporting Material S2.2).

## 5. Conclusion

The magnetic fields induced by the tDCS currents during fMRI studies can cause artifacts in the acquired EPI images that reach a similar strength as physiological BOLD activations. This may obscure real activations or yield false positive results. We identified underlying physical mechanisms and derived a simulation framework that enables assessing the severity of the artifacts and their impact on the fMRI results in dependence of the specific experimental design and analyses choices. We stringently validated the framework both in phantom experiments and measurements in humans. The simulation framework is publicly available to guide the design of tDCS-fMRI experiments that are robust to these artifacts.

## Data Availability

The simulation scripts and phantom dataset are publicly available at https://doi.org/10.17605/OSF.IO/GU6ER. Data of the human subjects cannot be shared publicly because of privacy restrictions.

## 6. Acknowledgements

This study was supported by the German Research Foundation (Research Unit 5429/1 (467143400), AT 1330/6-1, AT 1330/7-1, BL 977/4-1) and Lundbeck Foundation (grant R313-2019-622).

## References

[1] Antal A, Alekseichuk I, Bikson M, Brockmöller J, Brunoni AR, Chen R, et al. Low intensity transcranial electric stimulation: Safety, ethical, legal regulatory and application guidelines. Clin Neurophysiol 2017;128:1774–809. 10.1016/J.CLINPH.2017.06.001.

[2] Nitsche MA, Paulus W. Excitability changes induced in the human motor cortex by weak transcranial direct current stimulation. J Physiol 2000;527:633–9. 10.1111/j.1469-7793.2000.t01-1-00633.x.

[3] Stagg CJ, O’Shea J, Kincses ZT, Woolrich M, Matthews PM, Johansen-Berg H. Modulation of movement-associated cortical activation by transcranial direct current stimulation. Eur J Neurosci 2009;30:1412–23. 10.1111/j.1460-9568.2009.06937.x.

[4] Antal A, Polania R, Schmidt-Samoa C, Dechent P, Paulus W. Transcranial direct current stimulation over the primary motor cortex during fMRI. NeuroImage 2011;55:590–6. 10.1016/J.NEUROIMAGE.2010.11.085.

[5] Saiote C, Turi Z, Paulus W, Antal A. Combining functional magnetic resonance imaging with transcranial electrical stimulation. Front Hum Neurosci 2013. 10.3389/fnhum.2013.00435.

[6] Ekhtiari H, Ghobadi-Azbari P, Thielscher A, Antal A, Li LM, Shereen AD, et al. A checklist for assessing the methodological quality of concurrent tES-fMRI studies (ContES checklist): a consensus study and statement. Nat Protoc 2022;17:596–617. 10.1038/s41596-021-00664-5.

[7] Esmaeilpour Z, Shereen AD, Ghobadi-Azbari P, Datta A, Woods AJ, Ironside M, et al. Methodology for tDCS integration with fMRI. Hum Brain Mapp 2020;41:1950–67. 10.1002/hbm.24908.

[8] Antal A, Bikson M, Datta A, Lafon B, Dechent P, Parra LC, et al. Imaging artifacts induced by electrical stimulation during conventional fMRI of the brain. NeuroImage 2014;85:1040–7. 10.1016/j.neuroimage.2012.10.026.

[9] Gregersen F, Göksu C, Schaefers G, Xue R, Thielscher A, Hanson LG. Safety evaluation of a new setup for transcranial electric stimulation during magnetic resonance imaging. Brain Stimulat 2021;14:488–97. 10.1016/J.BRS.2021.02.019.

[10] Holland R, Leff AP, Josephs O, Galea JM, Desikan M, Price CJ, et al. Speech facilitation by left inferior frontal cortex stimulation. Curr Biol 2011;21:1403–7. 10.1016/j.cub.2011.07.021.

[11] Gbadeyan O, Steinhauser M, Mcmahon K,Meinzer M. Safety, Tolerability, Blinding Efficacy and Behavioural Effects of a Novel MRI-Compatible, High-Definition tDCS Set-Up. Brain Stimulat 2016;9:545–52. 10.1016/j.brs.2016.03.018.

[12] Meinzer M, Lindenberg R, Darkow R, Ulm L, Copland D, Flöel A. Transcranial Direct Current Stimulation and Simultaneous Functional Magnetic Resonance Imaging. J Vis Exp 2014:51730. 10.3791/51730.

[13] Jørgensen LM, Ohlhues Baandrup A, Mandeville J, Glud AN, Christian J, Sørensen H, et al. An fMRI-compatible system for targeted electrical stimulation. J Neurosci Methods 2022;378:109659. 10.1016/j.jneumeth.2022.109659.

[14] Weiskopf N, Josephs O, Ruff CC, Blankenburg F, Featherstone E, Thomas A, et al. Image Artifacts in Concurrent Transcranial Magnetic Stimulation (TMS) and fMRI Caused by Leakage Currents: Modeling and Compensation. J Magn Reson Imaging 2009;29:1211–7. 10.1002/jmri.21749.

[15] Jog M, Jann K, Yan L, Huang Y, Parra L, Narr K, et al. Concurrent Imaging of Markers of Current Flow and Neurophysiological Changes During tDCS. Front Neurosci 2020;14:374. 10.3389/fnins.2020.00374.

[16] Moeller S, Yacoub E, Olman CA, Auerbach E, Strupp J, Harel N, et al. Multiband multislice GE-EPI at 7 tesla, with 16-fold acceleration using partial parallel imaging with application to high spatial and temporal whole-brain fMRI. Magn Reson Med 2010;63:1144–53. 10.1002/MRM.22361.

[17] Grodzki DM, Jakob PM, Heismann B. Ultrashort echo time imaging using pointwise encoding time reduction with radial acquisition (PETRA). Magn Reson Med 2012;67:510–8. 10.1002/MRM.23017.

[18] Foerster BU, Tomasi D, Caparelli EC. Magnetic field shift due to mechanical vibration in functional magnetic resonance imaging. Magn Reson Med 2005;54:1261–7. 10.1002/mrm.20695.

[19] Kozlov M, Horner M, Kainz W, Weiskopf N, Möller HE. Modeling radio-frequency energy-induced heating due to the presence of transcranial electric stimulation setup at 3T. Magn Reson Mater Phys Biol Med 2020;33:793–807. 10.1007/s10334-020-00853-5.

[20] Bahr-Hosseini M, Bikson M. Neurovascular-modulation: A review of primary vascular responses to transcranial electrical stimulation as a mechanism of action. Brain Stimulat 2021;14:837–47. 10.1016/J.BRS.2021.04.015.

[21] Jenkinson M, Bannister P, Brady M, Smith S. Improved Optimization for the Robust and Accurate Linear Registration and Motion Correction of Brain Images. NeuroImage 2002;17:825–41. 10.1006/nimg.2002.1132.

[22] Jenkinson M, Beckmann CF, Behrens TEJ, Woolrich MW, Smith SM. FSL. NeuroImage 2012;62:782–90. 10.1016/J.NEUROIMAGE.2011.09.015.

[23] Thielscher A, Antunes A, Saturnino GB. Field modeling for transcranial magnetic stimulation: A useful tool to understand the physiological effects of TMS? Proc Annu Int Conf IEEE Eng Med Biol Soc EMBS 2015;2015-November:222–5. 10.1109/EMBC.2015.7318340.

[24] Jezzard P, Balaban RS. Correction for geometric distortion in echo planar images from B0 field variations. Magn Reson Med 1995;34:65–73. 10.1002/MRM.1910340111.

[25] Jezzard P, Clare S. Sources of distortion in functional MRI data. Hum Brain Mapp 1999;8:80–5. 10.1002/(SICI)1097-0193(1999)8:2/3%3C80::AID-HBM2%3E3.0.CO;2-C.

[26] Fuller WA. A Single Explanatory Variable. Meas. Error Models, John Wiley & Sons, Ltd; 1987, p. 1–99. 10.1002/9780470316665.ch1.

[27] Bossé M, Marland E, Rhoads G, Sanqui JA, BeMent Z. Generalizing R2 for deming regressions. Commun Stat - Theory Methods 2023;52:7731–43. 10.1080/03610926.2022.2059678.

[28] Bandettini PA, Wong EC, Hinks RS, Tikofsky RS, Hyde JS. Time course EPI of human brain function during task activation. Magn Reson Med 1992;25:390–7. 10.1002/mrm.1910250220.

[29] Zheng X, Alsop DC, Schlaug G. Effects of transcranial direct current stimulation (tDCS) on human regional cerebral blood flow. NeuroImage 2011;58:26–33. 10.1016/j.neuroimage.2011.06.018.

[30] Kwon YH, Ko MH, Ahn SH, Kim YH, Song JC, Lee CH, et al. Primary motor cortex activation by transcranial direct current stimulation in the human brain. Neurosci Lett 2008;435:56–9. 10.1016/J.NEULET.2008.02.012.

[31] Kwon YH, Jang SH. The enhanced cortical activation induced by transcranial direct current stimulation during hand movements. Neurosci Lett 2011;492:105–8. 10.1016/J.NEULET.2011.01.066.

[32] Liu ML, Karabanov AN, Piek M, Petersen ET, Thielscher A, Siebner HR. Short periods of bipolar anodal TDCS induce no instantaneous dose-dependent increase in cerebral blood flow in the targeted human motor cortex. Sci Rep 2022;12:9580. 10.1038/s41598-022-13091-7.

[33] Shinde AB, Lerud KD, Munsch F, Alsop DC, Schlaug G. Effects of tDCS dose and electrode montage on regional cerebral blood flow and motor behavior. NeuroImage 2021;237:118144. 10.1016/j.neuroimage.2021.118144.

[34] Shinde A, Mohapatra S, Schlaug G. Identifying the engagement of a brain network during a targeted tDCS-fMRI experiment using a machine learning approach. PLOS Comput Biol 2023;19:e1011012. 10.1371/journal.pcbi.1011012.

[35] Meinzer M, Antonenko D, Lindenberg R, Hetzer S, Ulm L, Avirame K, et al. Electrical Brain Stimulation Improves Cognitive Performance by Modulating Functional Connectivity and Task-Specific Activation. J Neurosci 2012;32:1859–66. 10.1523/JNEUROSCI.4812-11.2012.

[36] Göksu C, Hanson LG, Siebner HR, Ehses P, Scheffler K, Thielscher A. Human in-vivo brain magnetic resonance current density imaging (MRCDI). NeuroImage 2018;171:26–39. 10.1016/j.neuroimage.2017.12.075.

[37] Gregersen F, Eroğlu HH, Göksu C, Puonti O, Zuo Z, Thielscher A, et al. MR imaging of the magnetic fields induced by injected currents can guide improvements of individualized head volume conductor models. Imaging Neurosci 2024;2:1–15. 10.1162/IMAG_A_00176.

[38] Kasinadhuni AK, Indahlastari A, Chauhan M, Schär M, Mareci TH, Sadleir RJ. Imaging of current flow in the human head during transcranial electrical therapy. Brain Stimulat 2017;10:764–72. 10.1016/j.brs.2017.04.125.

