## Supplementary_Material for "Concurrent tDCS-fMRI: Impact of the current-induced magnetic fields on the measured BOLD signal"

### Supporting Information

#### Supporting Material S1: Geometric distortions caused by $\Delta \mathrm{B_{zc}}$

##### S1.1 Theory

Inhomogeneities of the static field $\mathrm{B_0}$ are known to cause image distortions [1]. The severity of these effects depends on the strength of the frequency-encoding (FE) gradients. In 2D EPI, the phase-encoding gradient blips act similarly to a weak constant FE gradient, which is the time average of the blips' moment during a full k-space traversal. Since this FE gradient is much weaker than the perpendicular readout gradient, the geometric distortions can, to a good approximation, be assumed to occur only in the blip direction (defined here as y). Furthermore, insignificant slice shifts are expected for broadband excitation. By taking the inverse Fourier Transform of the measured k-space signal along the perpendicular FE direction (x), the problem is reduced to a 1D imaging experiment with frequency encoding along y, for each location in x. Neglecting relaxation effects, the measured k-space is then given by [2]

$$S\Big(k(\tau)\Big) \propto \int_{\mathbb{R}} M_{\perp}(y) e^{-i\,\Delta\phi(y,\tau)} e^{-i\,2\pi\,k(\tau)\,y} dy\ , \tag{1}$$

where $k(\tau)$ is the k-space location sampled at an instant $\tau \in \left[-\frac{T_{ro}}{2}, \frac{T_{ro}}{2}\right]$ defined during a readout period of duration $T_{ro}$ and centered at the echo time $T_E$, where the imaging gradients are refocused (a centered readout is not important for the results, but it eases readability). Additionally, $\gamma$ is the gyromagnetic ratio of the imaged nucleus, $y$ is a coordinate in object space, $M_{\perp}$ is the complex transversal magnetization (weighed by the coil sensitivities) at $T_E$, and the complex exponentials $e^{-i\,\Delta\phi(y,\tau)}$ and $e^{-i\,2\pi\,k(\tau)\,y}$ represent the phases accrued due to $\mathrm{B_0}$ inhomogeneities and applied imaging gradients, respectively. The phase evolution of the recorded signal is slowed down due to the scanner demodulation at frequency $\omega_{ref}$. A constant $\mathrm{B_0}$ offset $\Delta B(y) = B_0(y) - \omega_{ref}/\gamma$ results in the phase term $\Delta\phi(y,\tau) = \gamma\,\Delta B(y)\,(T_E + \tau)$. Likewise, a constant readout gradient $G_{ro}$ preceded by a prewinder gradient with zero-th moment $-\frac{1}{2}\,G_{ro}\,T_{ro}$ results in the k-space trajectory $k(\tau) = \frac{\gamma}{2\pi}\,G_{ro}\,\tau$. Note the correspondence between $G_{ro}$ and the mean PE gradient in 2D EPI. Incorporating these considerations into Equation 1 results in

$$S\Big(k(\tau)\Big) \propto \int_{\mathbb{R}} M_{\perp}(y) e^{-i\,\gamma\,\Delta B(y)\,(T_E+\tau)} e^{-i\,\gamma\,G_{ro}\,\tau\,y} dy\ . \tag{2}$$

To improve readability, the time dependence in $k(\tau)$ will be omitted in the rest of the derivation.

So far, it has been assumed that the measured k-space is infinitely large. In practice, only a small fraction can be acquired due to time and signal decay constraints. Limiting the acquisition to $k \in [-\frac{K}{2}, \frac{K}{2}]$, $K \in \mathbb{R}$, results in the truncated k-space recording $S_{\text{rect}}(k) = S(k) \cdot \text{rect}\left(\frac{k}{K}\right)$, where $\text{rect}()$ is the rectangular function $\text{rect}\left(\frac{k}{K}\right) = \begin{cases} 1, \text{ if } |k| \leq K/2 \\ 0, \text{ if } |k| > K/2 \end{cases}$. Noting that $S(k)$ and $\text{rect}(\frac{k}{K})$ are proportional to the Fourier Transforms of $M_\perp(y) e^{-i\gamma \Delta B(y)(T_E+\tau)}$ and $K\text{sinc}(\pi K y)$, respectively, one can proceed by making use of the convolution theorem, which states that the Fourier Transform (FT) of the convolution of two functions is the product of their FTs:

$$S(k) \cdot \text{rect}\left(\frac{k}{K}\right) \propto \int_{\mathbb{R}} \left( \int_{\mathbb{R}} M_\perp(Y)\, e^{-i\gamma\, \Delta B(Y)\, (T_E+\tau)} \cdot \text{sinc}(K\pi(y-Y))\, dY \right) e^{-i\gamma\, G_{ro}\, \tau\, y}\, dy\ . \tag{3}$$

The term in parentheses represents the mixing of signals in the vicinity of $y$, which depends on the width of the $\text{sinc}$ point spread function (PSF) determined by the k-space coverage. Multiplying and dividing by a common factor, it can be rewritten as

$$\left( \int_{\mathbb{R}} M_\perp(Y)\, \text{sinc}(K\pi(y-Y))\, dY \right) \cdot \frac{\int_{\mathbb{R}} M_\perp(Y)\, \text{sinc}(K\pi(y-Y))\, e^{-i\gamma\, \Delta B(Y)\, (T_E+\tau)} dY}{\int_{\mathbb{R}} M_\perp(Y)\, \text{sinc}(K\pi(y-Y))\, dY}\ . \tag{4}$$

The integral on the left represents the mixing of the true magnetization (i.e., in the absence of field inhomogeneities) and will be denoted by $\overline{M_\perp}(y)$. The fraction on the right expresses the effects of the field offset $\Delta B$ as a complex weighted average of $e^{-i\gamma\, \Delta B(Y)\, (T_E+\tau)}$ over the $\text{sinc}$ PSF centered at $y$. It will be denoted by $\eta(y,\tau)\, e^{-i\gamma\, \overline{\Delta B}(y)\, (T_E+\tau)}$, where $\eta$ captures the signal loss due to the intravoxel dephasing caused by the spatial variation of $\Delta B$ within the $\text{sinc}$ kernel (see Supporting Material S4), and $\overline{\Delta B}$ is the apparent (i.e., measurable) mean field offset in the voxel centered in $y$. In tDCS-fMRI, $\overline{\Delta B}$ includes not only the field variations arising from magnetic susceptibility and shim differences within the sample, $\overline{B}_*$, but also the tiny magnetic fields induced by the tDCS currents, $\overline{B}_{zc}$. We can rewrite Equation 3 using these new definitions:

$$S_{\text{rect}}(k) \propto \int_{\mathbb{R}} \eta(y,\tau)\, \overline{M_\perp}(y)\, e^{-i\gamma \left[\overline{B}_*(y) + \overline{B}_{zc}(y)\right] (T_E+\tau)} e^{-i\gamma\, G_{ro}\, \tau\, y}\, dy\ . \tag{5}$$

The geometric distortions caused by $\overline{B}_*$ can be analyzed separately from those induced by the much weaker $\overline{B}_{zc}$:

$$S_{\text{rect}}(k) \propto \int_{\mathbb{R}} \eta(y,\tau)\, \overline{M_\perp}(y) e^{-i\gamma\, \overline{B}_*(y)\, T_E}\, e^{-i\gamma\, \overline{B}_{zc}(y)\, (T_E+\tau)}\, e^{-i\gamma\, G_{ro}\, \tau \left(y + \frac{\overline{B}_*(y)}{G_{ro}}\right)} dy\ . \tag{6}$$

The following change of variables and definitions can be made:

$$y' \equiv y + \frac{\overline{B}_*(y)}{G_{ro}} \equiv f(y) \tag{7a}$$

$$\frac{dy'}{dy} = 1 + \frac{\overline{G}_*(y)}{G_{ro}} \equiv Q_*(y)\ , \tag{7b}$$

where $y'$ is the coordinate, in the distorted image, assigned to a point in the undistorted image with coordinate $y$, and $\overline{G}_*(y) = d\overline{B}_*(y)/dy$ is the spatial gradient of the apparent field offset in the original (undistorted and blurred) space. The mapping between distorted and original coordinates, $y' = f(y)$, can be shown to be invertible if $\overline{G}_*(y)$ does not fully counteract $G_{ro}$. The quantity $Q_*(y)$ represents the local intensity change caused by spatial stretching/compression. Inserting these quantities into Equation 6 results in

$$S_{rect}(k) \propto \int_{\mathbb{R}} I_*(y')\, e^{-i\gamma \overline{B}_{zc}\left(f^{-1}(y')\right)(T_E+\tau)}\, e^{-i\gamma G_{ro}\tau y'} dy' \;. \tag{8}$$

Here, $I_*(y') \equiv \eta\left(f^{-1}(y'), \tau\right) \frac{\overline{M_\perp}\left(f^{-1}(y')\right)}{Q_*(f^{-1}(y'))}\, e^{-i\gamma \overline{B}_*\left(f^{-1}(y')\right) T_E}$ is the distorted image resulting from a simple inverse FT of $S_{rect}(k)$ in the absence of the current-induced magnetic fields $\overline{B}_{zc}$. $I_*(y')$ differs from the real (but blurred) magnetization $\overline{M_\perp}(y)$ by voxel displacements, magnitude scaling from both stretching/compression and intravoxel dephasing, and an additional phase variation. In the presence of $\overline{B}_{zc}$, the $\overline{B}_*$-distorted image $I_*$ will be further distorted, and the result can be derived in the same way as for the $\overline{B}_*$-related distortions. The key aspect here is that the $\overline{B}_*$-distorted current-induced magnetic fields, $\overline{B}_{zc}\left(f^{-1}(y')\right)$, should be used to describe those additional distortions in $I_*(y')$.

Equations 7a and 7b state that the amount of voxel shifting and intensity modulation is inversely proportional to the strength of the FE imaging gradients. In 2D EPI, a faster k-space traversal decreases the severity of these distortions by increasing the strength of the *mean* PE gradient blips, although this is limited by the readout bandwidth in the perpendicular FE direction. In-plane acceleration based on parallel imaging (PI) directly uses stronger gradient blips to undersample k-space in that direction. Both strategies are accompanied by a decrease in signal-to-noise ratio (SNR), but the PI-based approach suffers from an additional spatially-varying noise amplification determined by the receive coil geometry — the "g-factor" penalty [3]. Moreover, in-plane acceleration may compromise the performance of simultaneous multi-slice imaging by reducing the field of view in the PE direction, and thus the amount of PE shift that can be applied to each of the simultaneously acquired slices to improve their separation [4]. Hence, the tradeoff between geometric distortions and acceleration-related artifacts must be considered.

##### S1.2 Validation in a phantom

The theory derived here was validated in the phantom experiment depicted in Figure 3A&B. To ensure that only the local geometric distortions affected the real tDCS data, the global image shifts due to the online updating of the demodulation frequency (see Section S2) were avoided by disabling it during acquisition, and reconstructing the raw k-space data with our own software. In short, the $B_0$ drifts over time were estimated from the phase differences at the center of k-space between the two ghost correction lines acquired with the same readout gradient polarity. A general linear model (GLM) consisting of a fourth order polynomial and the current injection scheme was then fitted to the measurements, and only the fitted polynomial was used for frequency drift compensation. The estimated frequency offset in each EPI readout was corrected by applying an appropriate phase-roll in the PE direction to the k-space data, as derived in Section S2.1. This method will be denoted by “adapted frequency update”. Nyquist ghost correction was then performed using the 3-line navigator without PE blips following the method described in [5]. The 2D Inverse Fourier Transform was applied to the individual k-spaces, and the images obtained for each coil were finally combined using an adaptive combine algorithm that estimates smooth coil sensitivity maps from the coil-specific complex images [6, 7]. The obtained tDCS series was compared with the sham series reconstructed in the same way and in which the expected $\Delta B_{zc}$ distortions were artificially induced. The results are shown in Figure S1 for two example slices, demonstrating a very good correspondence between the simulated and measured artifacts.

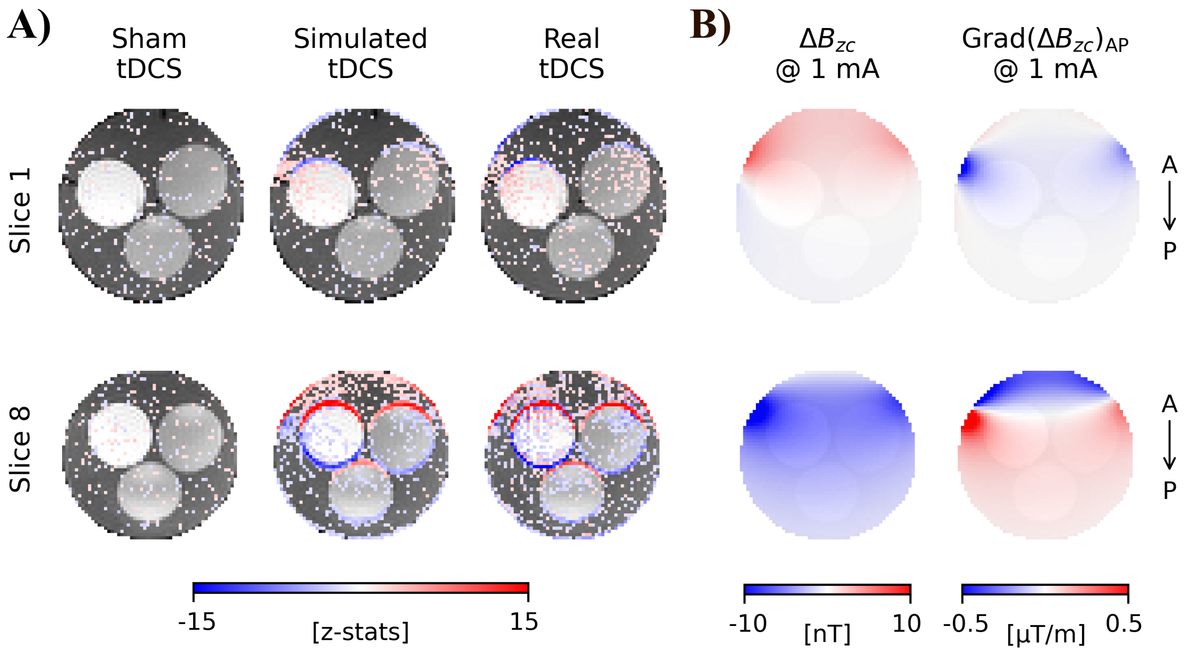


**Figure S1. Sensitivity of a first-level fMRI analysis to the EPI geometric distortions caused by the current-induced magnetic fields $\Delta B_{zc}$. A)** Activation maps (z-statistics thresholded at 1.65) for sham, simulated and real tDCS-fMRI data, for the slices indicated in Figure 3A. **B)** Simulated $\Delta B_{zc}$ (for a current intensity of 1 mA) and respective gradient in the phase-encoding direction, which was anterior (A) to posterior (P). The edge effects are mainly explained by the subvoxel shifts caused by $\Delta B_{zc}$, while the effects seen in homogeneous regions are essentially due to the stretching/compression described by its gradient.

#### Supporting Material S2: Impact of $\Delta B_{zc}$ on the demodulation frequency

##### S2.1 Theory

EPI sequences typically include an initial three-line navigator without phase-encoding (PE) blips for Nyquist ghost correction. These lines can also be used to estimate temporal drifts of the main field $B_0$ and the associated resonant frequency at the isocenter. An offset from the resonant frequency results in a phase difference between two measurements of the center of k-space at different echo times. Let $t_e$ and $t_e + 2\,t_{esp}$ be the echo times for the first and third correction lines, with $t_{esp}$ being the time between consecutive gradient echoes in EPI. A spatially-varying field offset $\Delta B(x, y, z)$ within the excited volume $V_{excit} \subset \mathbb{R}^3$ results in the measured signals $s_1$ and $s_3$ at those echo times:

$$s_1 \propto \int_{V_{excit}} M_\perp(x, y, z)\, e^{-i\gamma\,\Delta B(x,y,z)\,t_e}\, dV \tag{9a}$$

$$s_3 \propto \int_{V_{excit}} M_\perp(x, y, z)\, e^{-i\gamma\,\Delta B(x,y,z)\,(t_e+2t_{esp})}\, dV \tag{9b}$$

Here, $M_\perp$ is the complex transversal magnetization, the complex exponentials represent local phase terms induced by $\Delta B(x, y, z)$, and $dV = dx\,dy\,dz$. An average of $\Delta B$ over the excited volume can be estimated from the complex ratio between these two measurements:

$$\frac{s_3}{s_1} = \frac{\int_{V_{excit}} w(x, y, z)\, e^{-i\gamma\,\Delta B(x,y,z)\,2t_{esp}}\, dV}{\int_{V_{excit}} w(x, y, z)\, dV}\ , \tag{10}$$

with $w(x, y, z) \equiv M_\perp(x, y, z)\, e^{-i\gamma\,\Delta B(x,y,z)\,t_e}$. Equation 10 expresses the effects of the spatially-varying field offset $\Delta B(x, y, z)$ as a complex weighted average of the associated phase terms with weights $w(x, y, z)$. If the ghost correction lines are acquired shortly after excitation, $t_e$ is small and $w(x, y, z) \approx M_\perp(x, y, z)$. A first-order Taylor expansion of the complex exponential $e^{-i\gamma\,\Delta B(x,y,z)\,2t_{esp}}$, valid for a small phase accumulation during $t_{esp}$, results in a more intuitive expression of the phase difference between the centers of the two ghost correction lines:

$$\arg\left(\frac{s_3}{s_1}\right) \approx -2\gamma\, t_{esp} \frac{\int_{V_{excit}} M_\perp(x, y, z)\, \Delta B(x, y, z)\, dV}{\int_{V_{excit}} M_\perp(x, y, z) dV} \equiv -2\gamma\, t_{esp}\, \overline{\Delta B}_V\ , \tag{11}$$

where $\arg()$ is the argument of a complex number. The excited spins experience, on average, an off-resonance detuning that is approximately proportional to the weighted average of $\Delta B(x, y, z)$ over the excited volume, with the magnetization $M_\perp(x, y, z)$ as weights. This information can be used by the scanner to adjust the demodulation frequency, and the effect on the reconstructed image can easily be derived for the 1D experiment formulated in Equation 2. Adjusting the demodulation frequency by $\gamma\overline{\Delta B}_V$ results in the measured k-space signal

$$S(k) \propto \int_{\mathbb{R}} M_\perp(y) e^{-i\gamma\,\Delta B(y)\,(T_E+\tau)}\, e^{-i\gamma\,\overline{\Delta B}_V T_E}\, e^{-i\gamma\, G_{ro}\,\tau\left(y + \frac{\overline{\Delta B}_V}{G_{ro}}\right)} dy\ , \tag{12}$$

where all the remaining variables have the same meaning as in Equation 2. The lack of a spatial dependence of $\overline{\Delta B_V}$ results in much simpler geometric distortions: the change of variable $y' = y + \overline{\Delta B_V}/G_{ro}$ implies that all voxels are shifted by the same amount, which does not lead to additional stretching/compression nor the associated intensity modulation ($dy'/dy = 1$). Because the strength of the readout gradient $G_{ro}$ along a given direction determines the amount of shift in that direction, these occur almost exclusively along the PE direction for EPI imaging.

In the presence of current-induced magnetic fields, $B_{zc}$, the frequency update performed by the scanner will also compensate for the net frequency offset caused by the weighted average of $B_{zc}(x, y, z)$ over the excited volume. The negative side of it is the redistribution of part of the $B_{zc}$ effects throughout the imaged volume, leading for example to unexpected shifts in regions of weak $B_{zc}$ and reducing the expected ones in regions of strong $B_{zc}$.

##### S2.2 Validation in a phantom

The EPI sequence used in this study — "Multi-Band EPI BOLD C2P" from the Center for Magnetic Resonance Research (Minneapolis, Minnesota, USA) — offers the possibility to scan with or without online updating of the demodulation frequency. To validate the derived effects of $\Delta B_{zc}$ on the sample's average frequency (Equation 11), and understand how well these are mitigated by the online frequency updates implemented by this sequence, a current loop experiment was conducted in an FBIRN phantom (see Figure S2). The current "injection" scheme consisted of two main parts: (1) an initial period of alternating current polarity at the beginning of each repetition; (2) a final period with a much slower variation of the current intensity. Two almost identical acquisitions were performed, differing only in whether the online frequency updates were enabled. At each time point, the frequency offsets were estimated from the phase differences at the center of k-space between the two ghost correction lines acquired with the same readout gradient polarity (Figure S2C). The unprocessed measurements reveal the expected frequency drift over the course of the EPI time series when the online updates were disabled (in green). This series was then detrended to highlight the contribution of $\Delta B_{zc}$ to the center frequency offset. The measurements are well explained by the predictions based on the average of the simulated $\Delta B_{zc}$ over the imaged volume, weighed by the EPI image intensity (dashed black line). The online frequency updates (in blue) compensated for most of these offsets when the currents varied slowly over time, but did not succeed for rapidly varying currents. The implications of this for measurements of $\Delta B_{zc}$ based on the phase of the EPI images are discussed in Section S3.

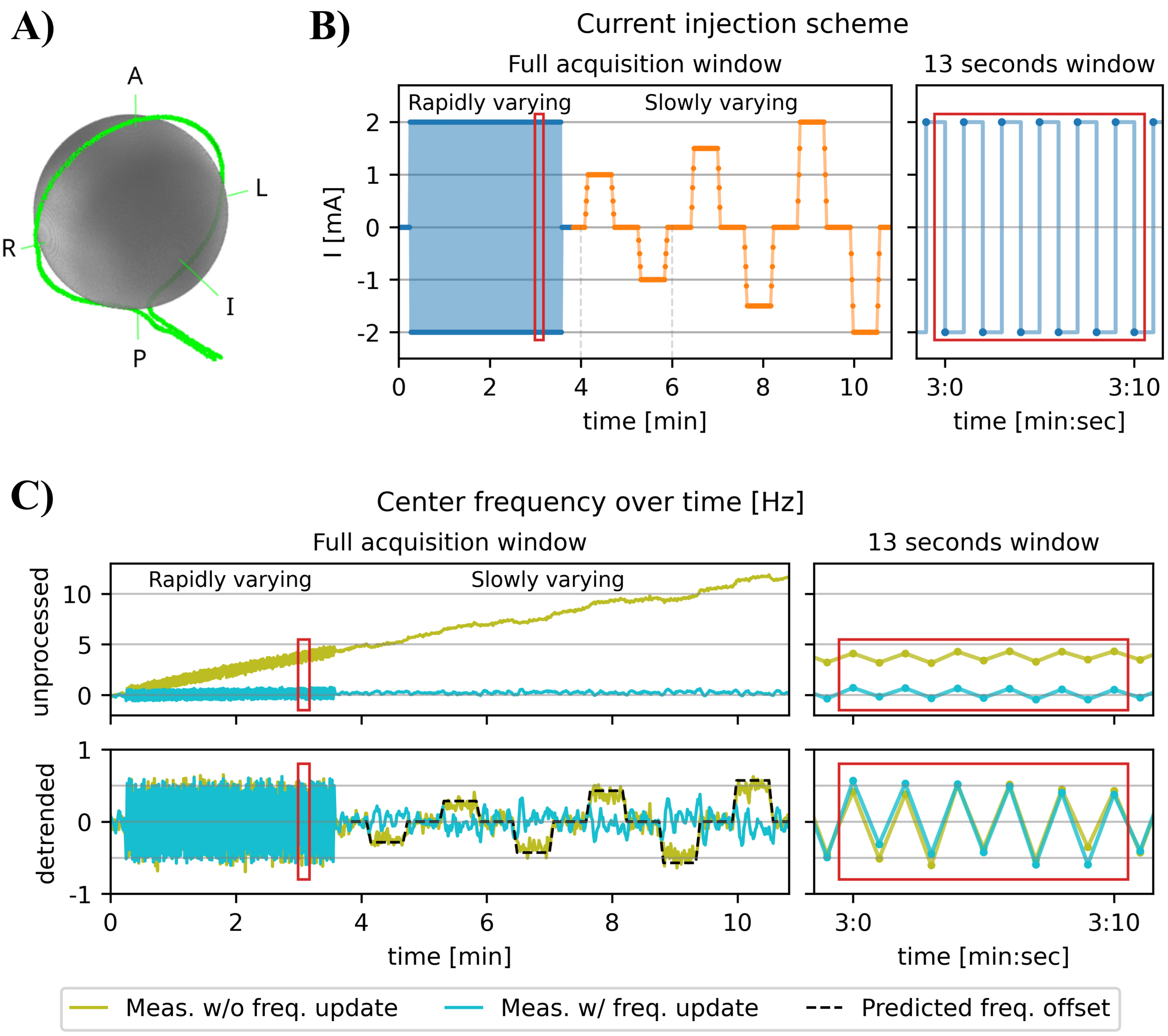


**Figure S2.** Effect of the current-induced magnetic fields $\Delta B_{zc}$ on the scanner's center frequency. **A)** 3D PETRA image of the FBIRN phantom, with the manually tracked cable loop shown in green. **B)** Current "injection" scheme. A zoomed version of the rapidly varying part is shown on the right for the window indicated by the red rectangle. **C)** Time series of the center frequency offset in acquisitions with and without online frequency updates. The unprocessed measurements (top) reveal the expected frequency drift over the course of the EPI time series when the online updates were disabled. The detrended series (bottom) shows a clear contribution of $\Delta B_{zc}$ to the sample's average frequency offset, and how this is partially mitigated by the mentioned updates.

A second loop experiment was performed to confirm that the implemented artifact simulation pipeline can be used to predict data acquired either with or without demodulation frequency updates. For this specific sequence, disabling the online updates automatically triggers a correction of the resulting image drifts in the PE direction during post-processing — this option will be referred to as "offline frequency update". To better assess the effects of these global image shifts on the fMRI analyses, the experiment was conducted in an in-house-built phantom with regions of high $T_2^*$ contrast (cf. Figure 2A&B). Measurements with online and offline frequency updates are compared in Figure S3. It appears that these have almost identical effects on the magnitude images, as judged by the remarkable similarity between the results of first level fMRI analyses of the current injection scheme. Note that these differ from the results shown in Figure 2F, where the images were reconstructed from raw k-space data using in-house-developed software, which applied the adapted frequency drift compensation described in Section S1.2.

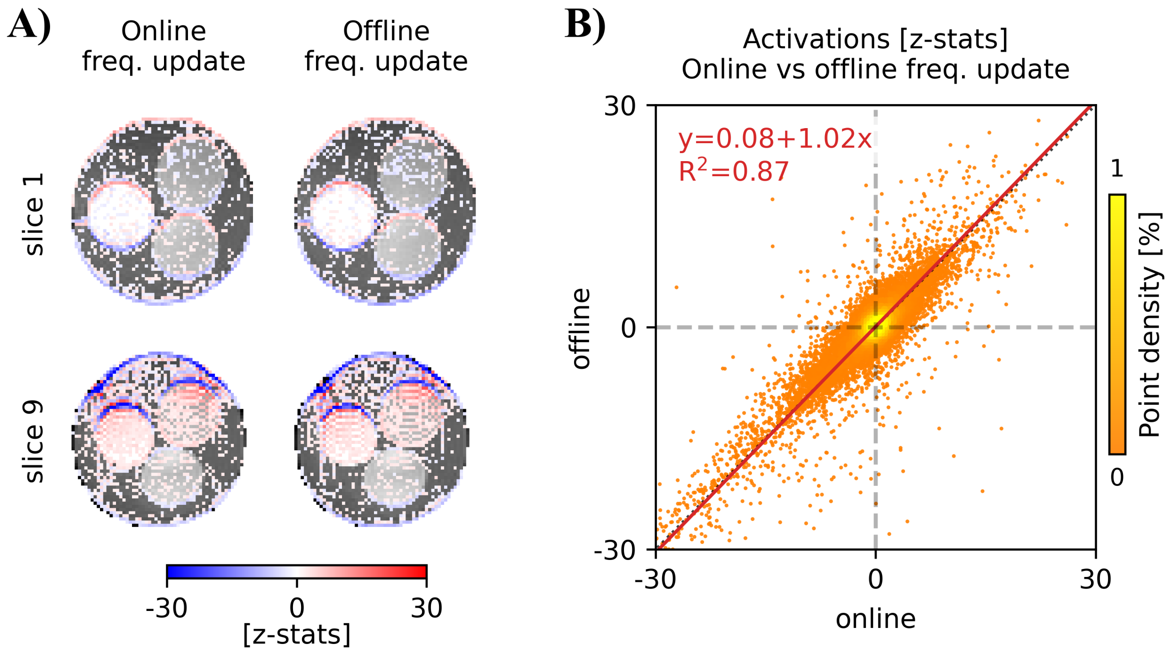


**Figure S3. Comparison of the $\Delta B_{zc}$ effects on fMRI series acquired with online and offline updating of the center frequency. A)** Activation maps (z-statistics thresholded at 1.65) from a first-level analysis of the fMRI data with online and offline frequency update, for the slices indicated in Figure 2B. The similarity between the two is remarkable. **B)** Scatter plot of the non-thresholded z-statistics for all slices of the fMRI series acquired with online vs offline frequency update. The linear fit depicted in red supports the equivalence between the two acquisition strategies when using the default reconstruction software.

#### Supporting Material S3: Measurements and simulations of $\Delta \mathrm{B}_{\mathrm{zc}}$

Biot-Savart-based simulations of $\Delta B_{zc}$, computed along known current paths, have been used as ground truth for $\Delta B_{zc}$ measurements in MRCDI [8–11]. Here, the magnetic fields in the brain were approximated by simulations based on the cable paths (manually tracked in PETRA images). This is a good approximation, since the magnetic fields from the currents in the electrode cables are much larger than those from the currents in the head (see Figure 1C). Nevertheless, it is still important to know whether the fields induced by block-design tDCS can be accurately determined based on the perturbations that they cause in the phase of the EPI images. This may be useful, for example, when images of the cable paths are not available. In addition, such a comparison serves as an additional validation of the Biot-Savart simulations.

This investigation was part of the experiment described in Figure S2A&B. The first half of the acquisition consisted of current injection with alternating polarity in synchrony with the sequence, as is typically done in MRCDI [8, 10–12]); the second part consisted of currents applied in a balanced block design (each block including 30 seconds of constant current, surrounded by 5 seconds of ramp-up and ramp-down), as often done in tDCS [13, 14]. Identical measurements were performed with and without online frequency drift correction, and reconstructed using the default software associated with this specific sequence (see Section S2.2). Voxel-wise temporal unwrapping of the phase time-series was performed [15], and general linear models consisting of a third order polynomial and the current injection scheme were independently fit to the first and second parts of the acquisition. Finally, the measured magnetic fields were corrected for the geometric distortions from the much stronger static $B_0$-field inhomogeneities using a separately acquired GRE field map, according to [11]. The simulated $\Delta B_{zc}$ for 1-mA currents in the cable loop was compared to the $\Delta B_{zc}$ per mA measured in each of the four conditions tested (fast and slow current variation, each with online and offline frequency correction). The accuracy of the measurements was quantitatively assessed through an ordinary least squares linear regression between the simulated and measured $\Delta B_{zc}$, with the noise-free simulations as independent variable. The results shown in Figure S4 indicate a high similarity between the simulation and measurements, except when the latter were based on the slowly varying period in the acquisition with frequency update. The online adjustments of the demodulation frequency "slow down" the phase accumulation due to the current-induced magnetic fields, resulting in an underestimation of these fields. The measured field offset of 7.25 nT is well explained by the frequency adjustments shown in Figure S2C: At 1 mA, $\Delta B_{zc}$ shifted the measured center frequency by approximately 0.3 Hz (in green), which corresponds to an approximate field offset of 7.05 nT. The dynamic updates of the demodulation frequency compensated for this (in blue), leading to an underestimation of $\Delta B_{zc}$ by approximately the same amount. Therefore, the slowly-varying magnetic fields induced by block-design tDCS can only be determined up to an additive constant using this specific sequence and the standard online frequency updates. Interestingly, the equivalence between the online and offline updating approaches reported in section S2.2 for the magnitude images is not observed in the phase images: The offline method only corrects for the image shifts, leaving the temporal drifts in the phase images intact. MRCDI measurements based on this sequence [10, 11] can still be performed with the default online approach, given its inability to compensate for the rapidly-varying magnetic fields.

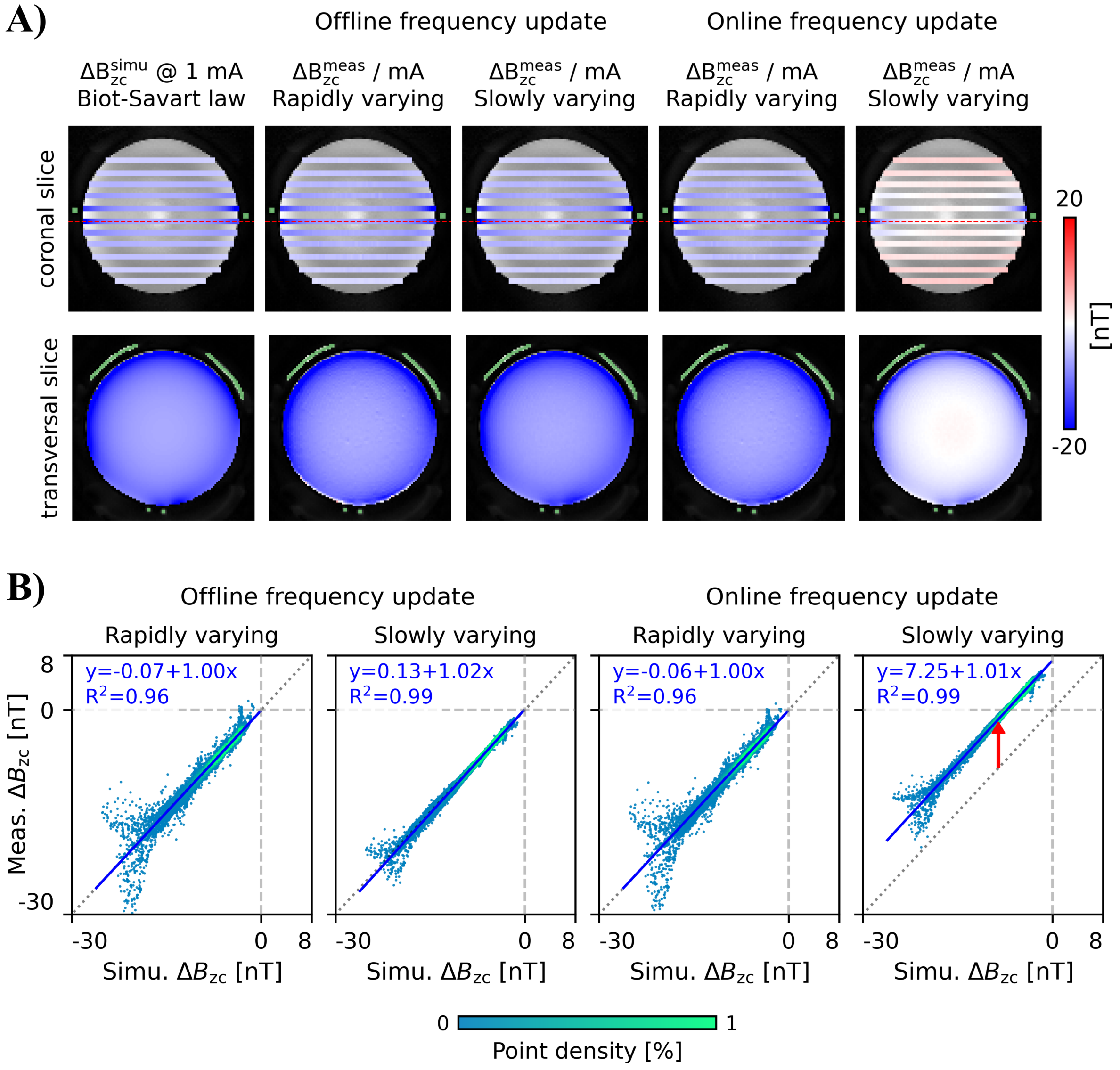


**Figure S4. Comparison between a simulation of the current-induced magnetic fields based on the tracked cable path ($\Delta \mathrm{B}_{\mathrm{zc}}^{\mathrm{simu}}$) and measurements based on the phase perturbations in the EPI images ($\Delta \mathrm{B}_{\mathrm{zc}}^{\mathrm{meas}}$).** The experimental setup is shown in Figures S2A&B. **A)** Simulated and measured $\Delta B_{zc}$ per mA overlaid on the Petra image, with the cable loop indicated in green. Results are shown for a coronal slice and the transversal slice indicated by the dashed red line. Measurements were performed with online and offline center frequency updates, and the two current injection periods were analyzed separately. Note the high similarity between simulation and measurements, except those based on the slowly varying period in the acquisition with online frequency update. **B)** Scatter plot and linear fit between the simulated and measured $\Delta B_{zc}$, for all measured slices. The red arrow in the right-most plot indicates the erroneous field offset in the measurements based on the slowly varying period in the acquisition with online frequency update.

#### Supporting Material S4: Intravoxel dephasing caused by $\Delta \mathrm{B_{zc}}$

Equation 3 models the signal mixing that results from a limited k-space coverage, and the exact result is given in Equation 4. Although the voxel size $\delta$ is defined as the full width at half maximum (FWHM) of the $\mathrm{sinc}$ PSF, the signal in each voxel has a substantial contribution from transversal magnetization outside that interval. For simplicity, only the center and first side lobes will be considered here, since they account for approximately 95% of the total $\mathrm{sinc}$'s power. Thus, assuming a constant magnetization $M_\perp(Y) \approx M_\perp(y)$ and a linear field offset $\Delta B(Y) \approx \Delta B(y) + G_y \cdot (Y - y)$ in an interval of width $\frac{4}{K}$ centered in $y$, the convolution integral in parentheses in Equation 3 is approximated by

$$M_\perp(y)\, e^{-i\gamma\,\Delta B(y)\,t} \int_{y-\frac{2}{K}}^{y+\frac{2}{K}} e^{-i\gamma\, G_y\cdot(Y-y)\,t} \cdot \mathrm{sinc}\left(K\pi(Y-y)\right)\, dY\ , \tag{13}$$

where $t = T_E + \tau$ and $G_y$ is the y-gradient of the field inhomogeneity at location $y$. With the change of variables $Y' = K\pi(Y - y)$, the integral in Equation 13 becomes

$$\frac{1}{K\pi}\int_{-2\pi}^{2\pi} e^{-i\,\frac{\gamma\, G_y\, t}{K\pi}Y'} \cdot \mathrm{sinc}(Y')\, dY' \propto \int_{0}^{2\pi} \cos\left(\frac{\gamma\, G_y\, t}{K\pi}Y'\right) \cdot \mathrm{sinc}(Y')\, dY'\ . \tag{14}$$

The right-hand side of Equation 14 results from writing $e^{-i\,\frac{\gamma\, G_y\, t}{K\pi}Y'}$ in cartesian form and exploiting the parity of the resulting functions, given the symmetry of the integration domain around 0. Noting that the FWHM of $\mathrm{sinc}\left(K\pi(Y-y)\right)$ is approximately $\frac{1.2}{K}$, the voxel size can be expressed as $\delta \approx \frac{4}{K\pi}$ such that the expected signal change can be written more intuitively as a function of the intravoxel field variation $G_y\,\delta$. In a worst-case scenario, the weak and spatially smooth magnetic fields induced by the tDCS currents ($\leq$ 4 mA) may vary up to 1-2 nT across a voxel (close to where the cables bend). For the echo times typically used in fMRI (30-40 ms), the expected signal variation due to intravoxel dephasing is of the order of $10^{-5}$, which is three orders of magnitude lower than that estimated due to geometric distortions (see, for example, Figure 1C). Hence, it can be safely ignored. This conclusion is unchanged for current-induced field variation over relevant slice profiles.

#### Supporting Material S5: Sensitivity of rigid motion correction to $\Delta B_{zc}$

The sensitivity of rigid motion correction methods to the $\Delta B_{zc}$-induced artifacts was evaluated in the phantom experiment described in Figure 3A&B. The current injection scheme was clearly represented in three of the six motion parameters estimated using *realign* from SPM12 [16], indicating that the $\Delta B_{zc}$–induced artifacts were perceived as movement by the registration algorithm (Figure S5A). Applying the complete motion parameters (termed "standard motion correction" here) changed the activation patterns in a non-trivial way (Figure S5B): In an attempt to correct for the local and non-linear $\Delta B_{zc}$–induced geometric distortions, global rotations and translations of the object (linear operations) succeeded in mitigating some of those distortions but at the cost of inducing unexpected effects elsewhere. An "adapted motion correction" pipeline was implemented, where a general linear model consisting of a sixth order polynomial and the current injection scheme was fitted to each individual motion parameter (black lines in Figure S5A), followed by subtraction of the fitted current regressors from the original parameters. The adapted parameters were then applied to the image series using *reslice* from SPM12. This method did not change the well-understood and predictable activation patterns observed without correction (Figure S5B).

In humans, the impact of the currents on the motion parameters was unclear due to physiological fluctuations of much larger amplitude (e.g., real movement, breathing). Nevertheless, even if relevant in some cases, these effects are expected to be present in the real and simulated tDCS data to a similar extent. Therefore, we used the standard correction performed by FSL's *MCFLIRT* [17] to obtain the results presented in Figure 5. For completeness, we report here the equivalent results obtained with the standard and adapted motion corrections based on SPM12 described above (Figures S6 and S7, respectively). As expected, the three methods provided comparable results.

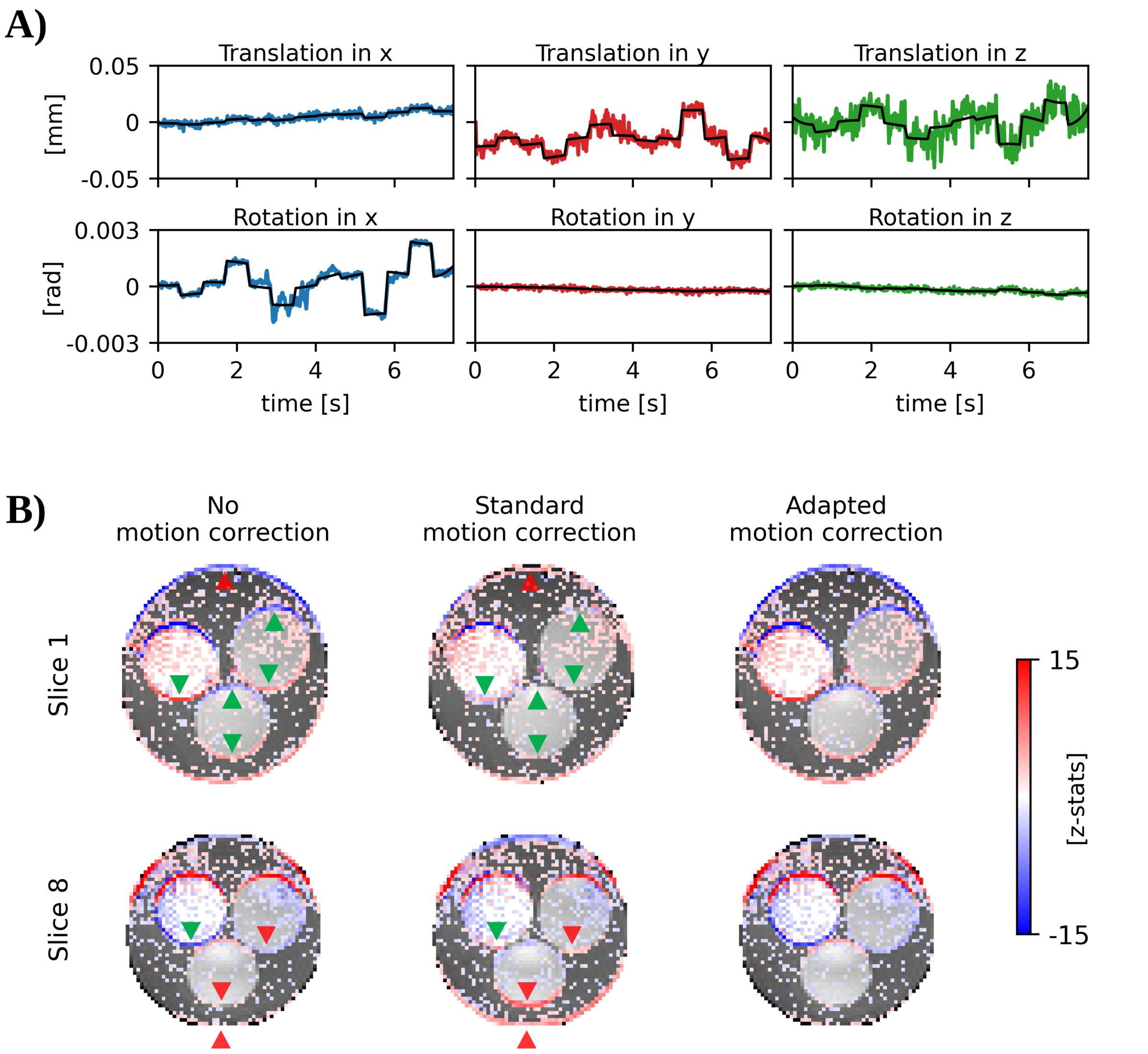


**Figure S5. Sensitivity of rigid motion correction to the current-induced magnetic fields ($\Delta B_{zc}$).** **A)** Motion parameters estimated by SPM12's *realign* algorithm with 6 degrees of freedom. **B)** Activation maps (z-statistics thresholded at 1.65) from first-level analyses of real tDCS-fMRI data with no, standard and adapted motion correction, for two example slices. The applied rotations and translations mitigated some of those activations (green arrows), but induced unexpected ones elsewhere (red arrows).

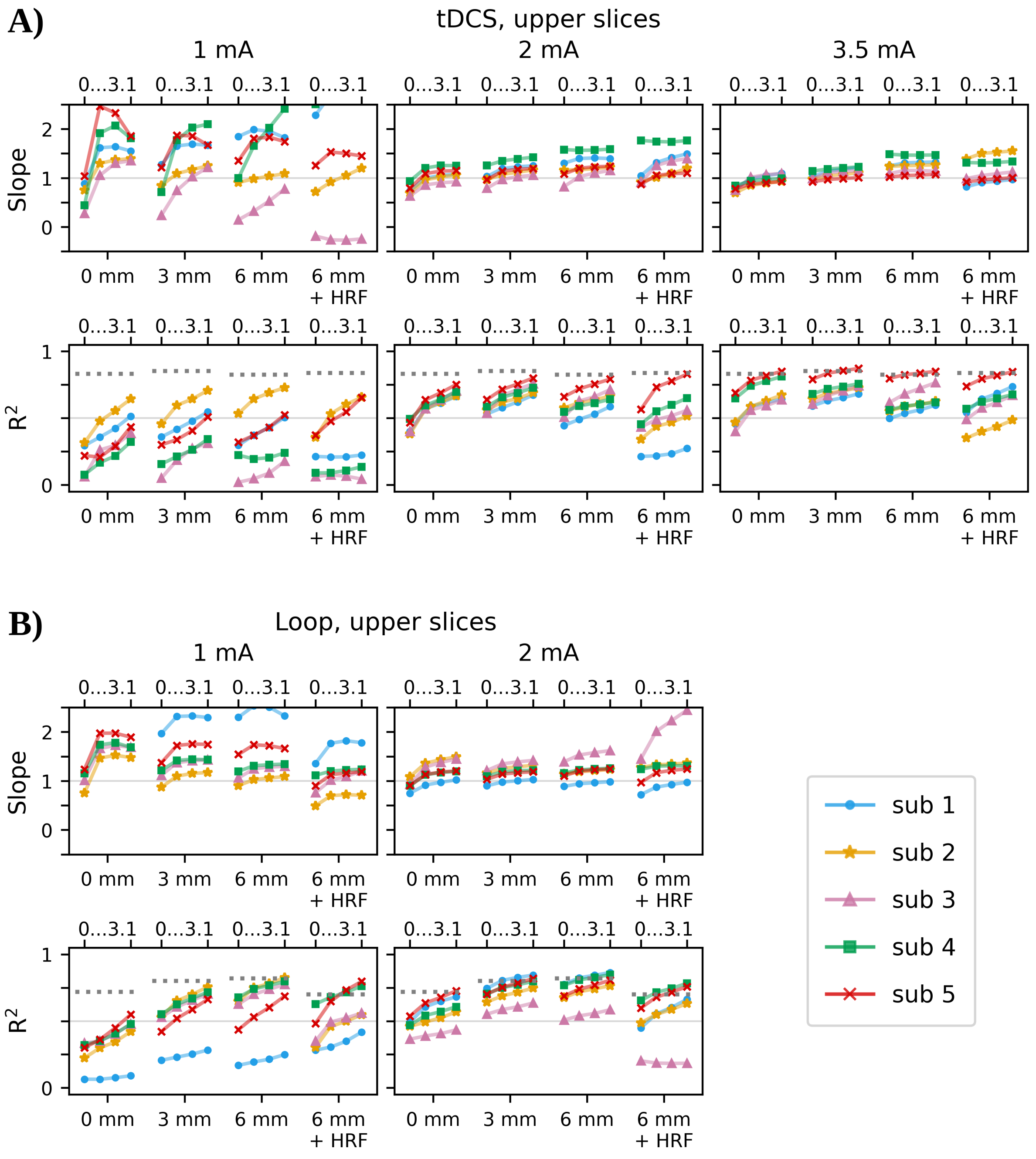


**Figure S6. Summary results for all subjects and tested current intensities in the *upper* half of the FoV, for a *standard* rigid-body motion correction using SPM12**. Shown are the slopes and coefficients of determination ($R^2$) of the linear fits to the z-statistics calculated for real and simulated current-induced artifacts, for spatial smoothing levels of the EPI time series varied between 0, 3 and 6 mm FWHM, as indicated on the horizontal axes. The time series smoothed with 6 mm were additionally analyzed using a version of the current injection scheme that was convolved with the Hemodynamic Response Function (HRF), replicating a standard approach for BOLD fMRI analyses. The linear fits were evaluated for different thresholding levels (0, 1.65, 2.3 and 3.1) of the simulated data (indicated by 0...3.1 on the upper horizontal axes). The approximate upper bounds for $R^2$ due to measurement noise are shown as horizontal dotted lines. **A)** tDCS results for 1, 2 and 3.5 mA. **B)** Loop results for 1 and 2 mA.

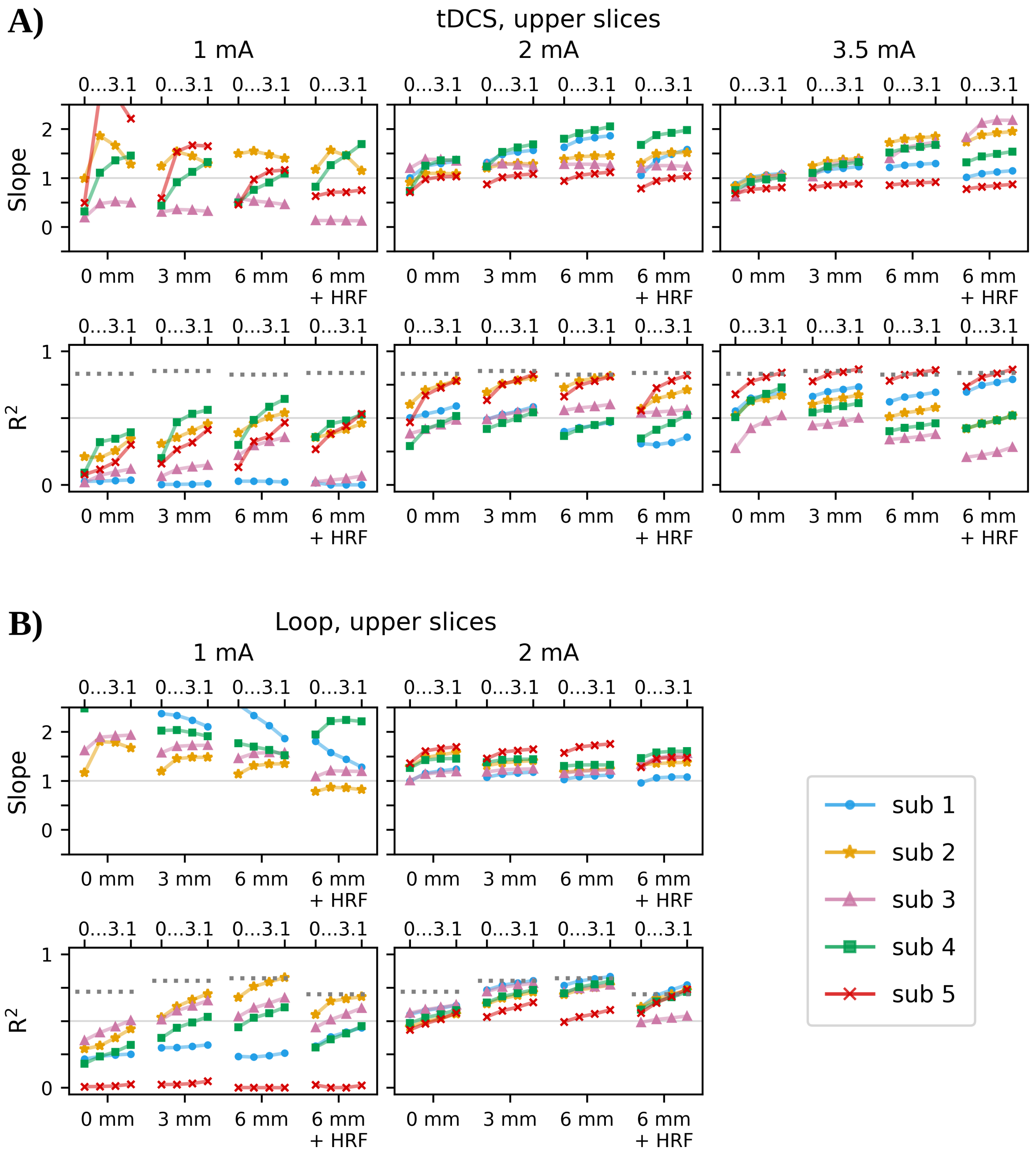


**Figure S7. Summary results for all subjects and tested current intensities in the *upper* half of the FoV, for a *adapted* rigid-body motion correction based on SPM12**. Shown are the slopes and coefficients of determination ($R^2$) of the linear fits to the z-statistics calculated for real and simulated current-induced artifacts, for spatial smoothing levels of the EPI time series varied between 0, 3 and 6 mm FWHM, as indicated on the horizontal axes. The time series smoothed with 6 mm were additionally analyzed using a version of the current injection scheme that was convolved with the Hemodynamic Response Function (HRF), replicating a standard approach for BOLD fMRI analyses. The linear fits were evaluated for different thresholding levels (0, 1.65, 2.3 and 3.1) of the simulated data (0...3.1 on the upper horizontal axes). The approximate upper bounds for $R^2$ due to measurement noise are shown as horizontal dotted lines. **A)** tDCS results for 1, 2 and 3.5 mA. **B)** Loop results for 1 and 2 mA.

#### Supporting Material S6: Validation of simultaneous multislice acquisitions

A conservative multiband factor of 3 was used in the main study to avoid interslice leakage artifacts [4, 18], which could camouflage or mix the effects of $\Delta B_{zc}$ between the simultaneously acquired slices. To confirm the negligible impact of this moderate acceleration on the $\Delta B_{zc}$-induced "activation" patterns, single-slice acquisitions were also performed in three subjects for real tDCS and loop with 3.5 and 2 mA currents, respectively. Moreover, these measurements allowed us to estimate the maximum expected coefficient of determination ($R^2$) of the linear fits to the simulated versus real activation maps, given the measurement noise.

To keep the same acquisition time (and thus stimulation paradigm), only 16 slices could be sequentially acquired within the chosen $T_R$, and an interslice gap of 1 mm was imposed for increased brain coverage. The corresponding multi-slice measurements were interpolated at the center of the individually acquired slices for a voxel-wise comparison of the activations determined with each acquisition strategy. A standard rigid-body motion correction was performed on both series using FSL MCFLIRT, with the first time point of the multi-slice acquisition used as the reference volume. First-level fMRI analyses were conducted as described in Section 2.4 of the main text, and linear Deming regression was performed on the activations obtained

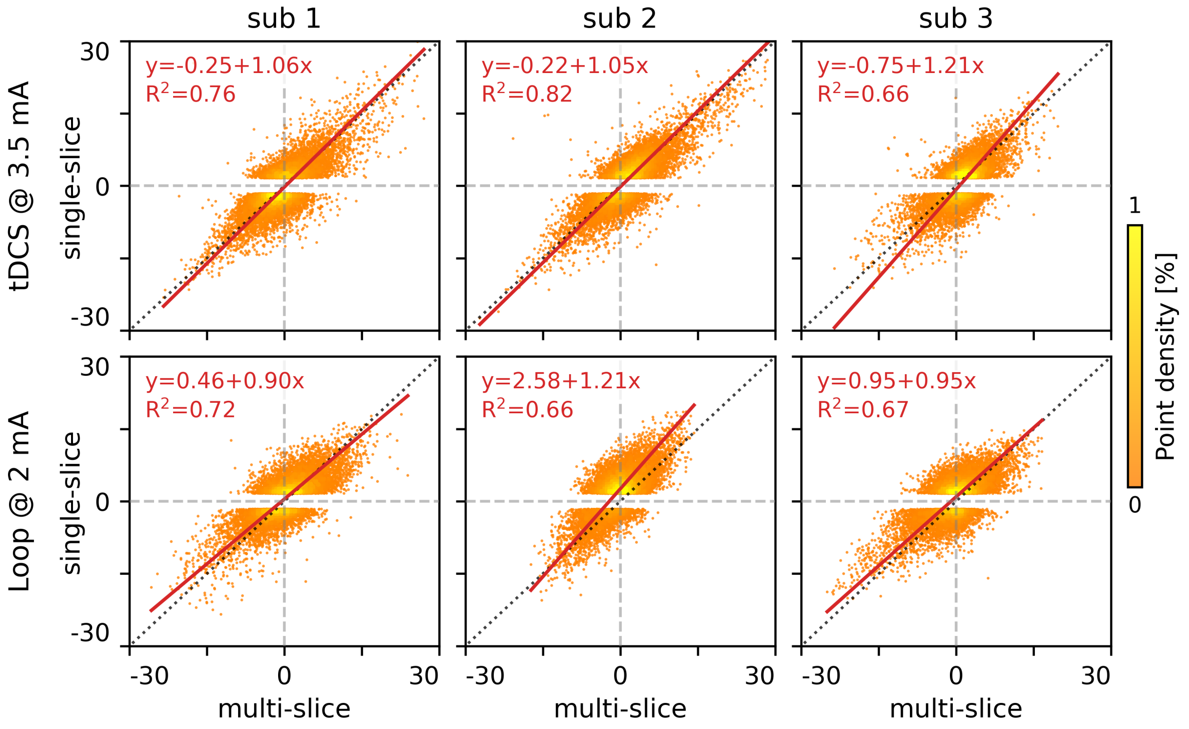


**Figure S8. Scatter plots and linear fits of the activations determined for single-slice vs simultaneous multi-slice measurements.** A spatial smoothing of 3 mm FWHM was applied to the image series before conducting the fMRI analyses. The linear fits to the activation maps are depicted by red lines and the respective parameters annotated in the upper left corners. To ensure consistency with the results shown in Figures 4 and S10, a threshold of 1.65 was applied to the ordinate data (single-slice activations on the vertical axis) prior to the fit.

for single- versus multi-slice measurements. Figure S8 shows the results obtained for a spatial smoothing of 3 mm FWHM. For consistency with Figure 5, a threshold of 1.65 was applied to the single-slice activations prior to the fit. The slopes and $R^2$ values computed for different thresholds and smoothing levels are shown in Figure S9.

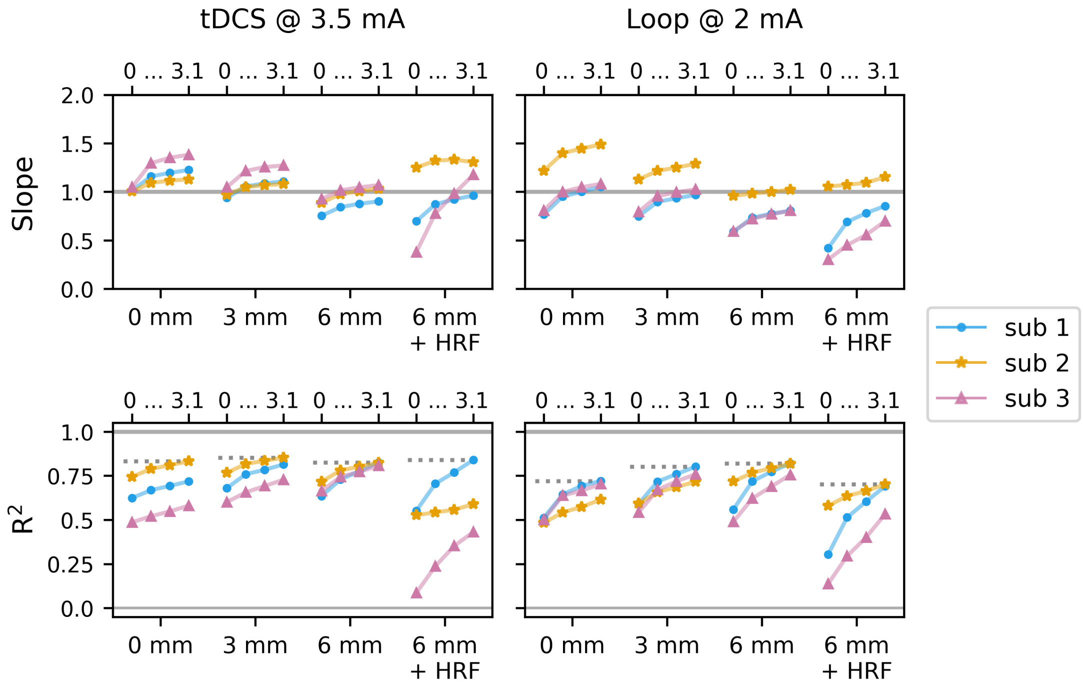


**Figure S9. Slopes and coefficients of determination ($R^2$) of the linear fits to the z-statistics obtained with single- vs multi-slice measurements.** Shown are the results for different smoothing levels applied to the time series (0, 3 and 6 mm FWHM; the latter was additionally analyzed using a version of the current injection scheme that was convolved with the Hemodynamic Response Function, HRF), as indicated on the lower horizontal axes. The thresholds applied to the single-slice data (0, 1.65, 2.3 and 3.1) are indicated on the upper horizontal axes. Thresholding was applied to the single-slice activations to make the results shown here comparable to those illustrated in Figures 5, S6, S7, S11 and S12. For each smoothing level, the maximum $R^2$ values obtained are indicated as horizontal dotted lines, giving an estimate of the maximally achievable $R^2$ in the fits in Figures 5, S6, S7 and S12.

#### Supporting Material S7: Artifact simulation — additional figures

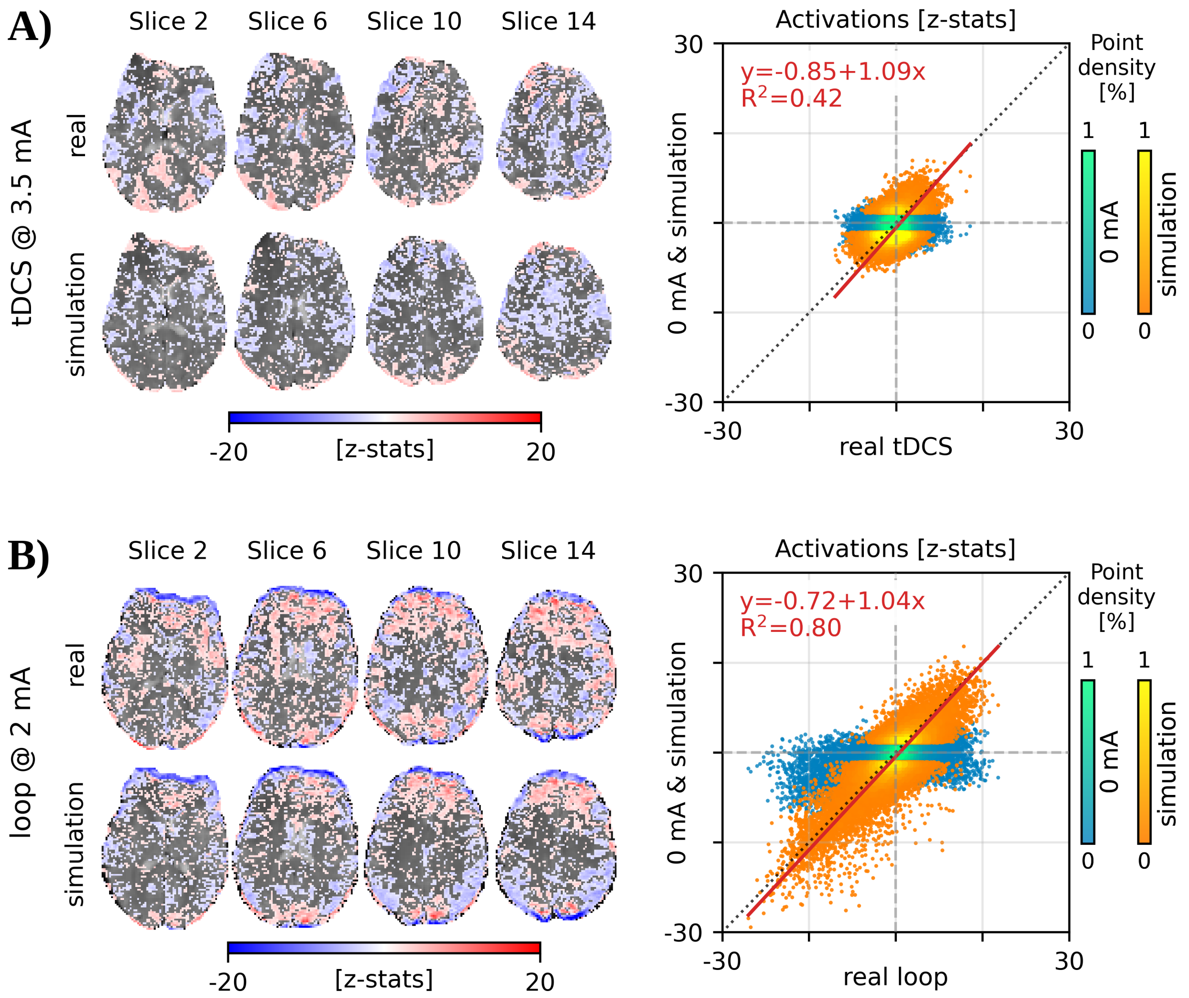


**Figure S10. Simulation of the artifacts caused by the current-induced magnetic fields ($\Delta \mathbf{B_{zc}}$) in the *lower* half of the EPI field of view (FoV), complementing Figure 4**. The simulated $\Delta B_{zc}$ for 3.5 mA and 2 mA currents flowing in the electrode leads and cable loop, respectively, are shown in Figure 4A. **A/B) Left:** Activation maps (z-statistics thresholded at 1.6) for the real and simulated tDCS/loop in the four example slices indicated by the dashed cyan lines in Figure 4A. The data is overlaid on the first image of the corresponding fMRI series. **Right:** Scatter plot of the activations obtained for 0 mA and simulated tDCS/loop against real tDCS/loop, and linear fit between the simulated and real activations. Please note that real tDCS resulted in much weaker remote effects than the cable loop in the lower slices distant to the cable paths shown here. The z-statistics of the simulated tDCS/loop were thresholded at 1.65 before computing the linear fit between real and simulated activations.

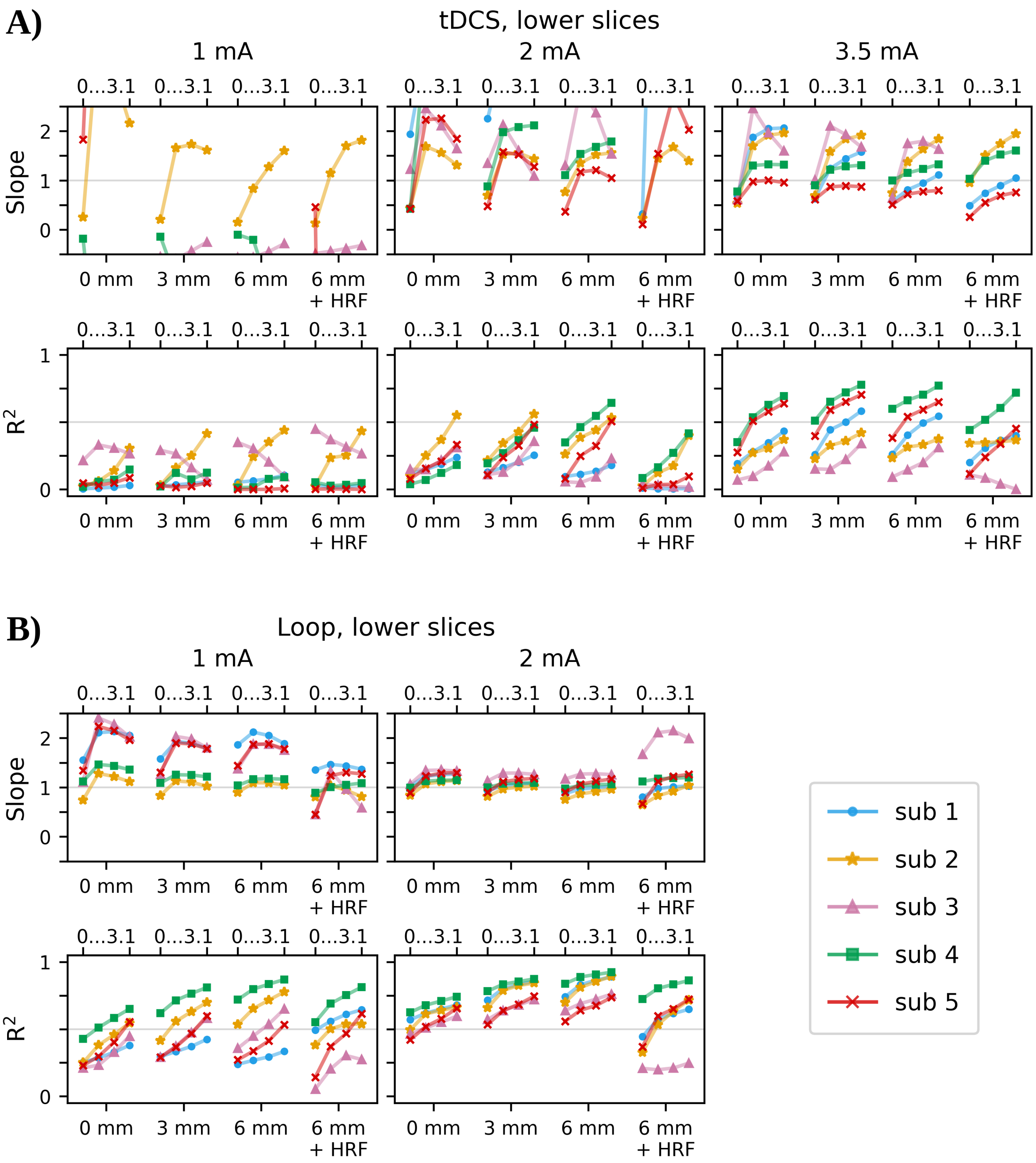


**Figure S11. Summary results for all subjects and tested current intensities in the *lower* half of the FoV, complementing Figure 5**. The preprocessing pipeline included a *standard* rigid-body motion correction using MCFLIRT. Shown are the slopes and coefficients of determination ($R^2$) of linear fits to the z-statistics calculated for real and simulated artifacts, for spatial smoothing levels of the EPI time series varied between 0, 3 and 6 mm FWHM, as indicated on the horizontal axes. The time series smoothed with 6 mm were additionally analyzed using a version of the current injection scheme that was convolved with the Hemodynamic Response Function (HRF), replicating a standard approach for BOLD fMRI analyses. The linear fits were evaluated for different thresholding levels (0, 1.65, 2.3 and 3.1) of the simulated data (indicated by 0...3.1 on the upper horizontal axes). **A)** tDCS results for 1, 2 and 3.5 mA. In line with Figure S10, the lead configuration for real tDCS resulted in weak remote effects, so that correlations between simulated and measured effects remained lower than for the upper slices even at high current intensities. **B)** Loop results for 1 and 2 mA. The loop configuration caused stronger remote effects, which is apparent from the good correlations at 2 mA for all participants.

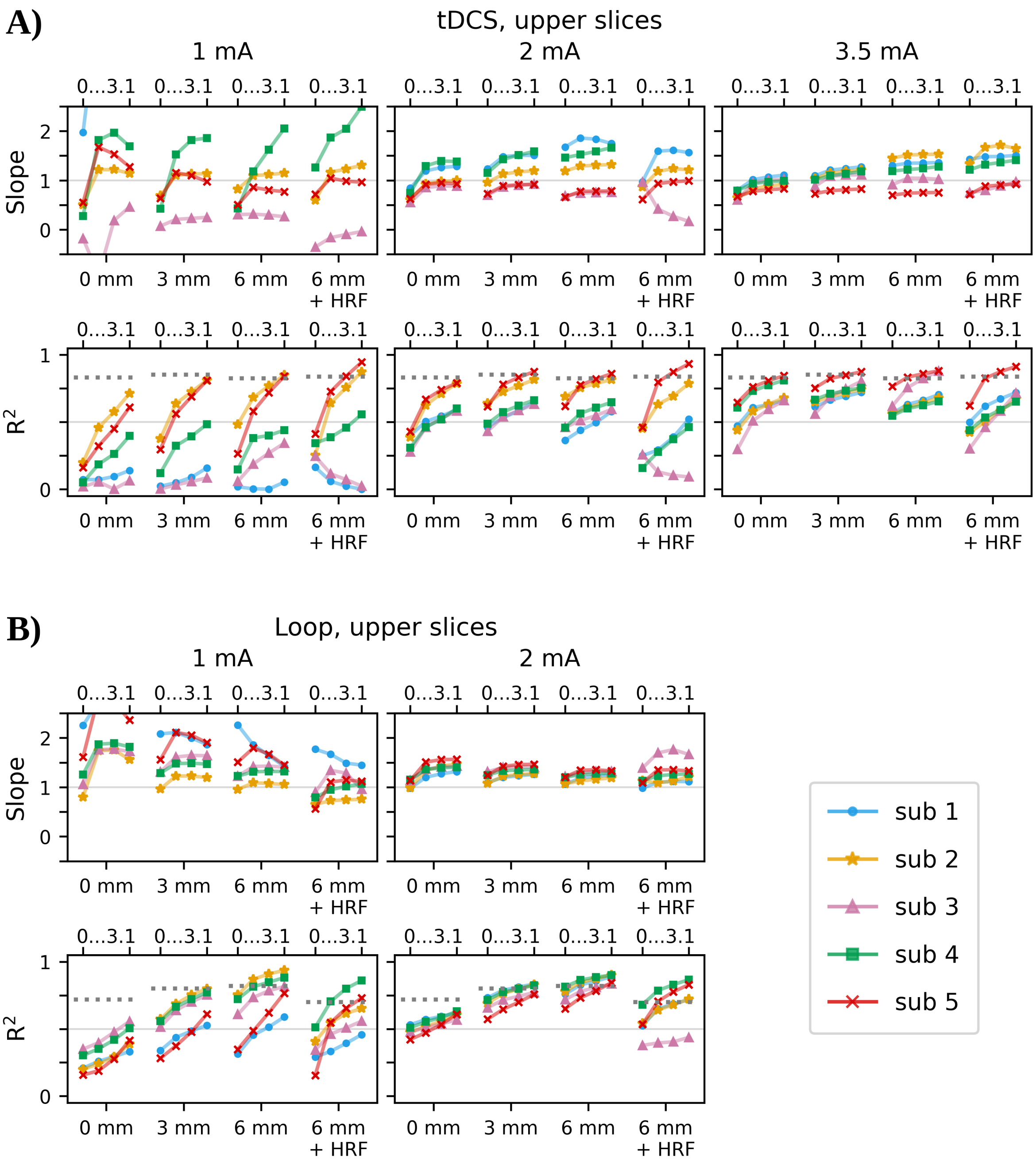


**Figure S12. Summary results for all subjects and tested current intensities in the *upper* half of the FoV, for fMRI analyses performed *with* prewhitening**. The input data to FSL FEAT was the same as for Figure 5. Shown are the slopes and coefficients of determination ($R^2$) of linear fits to the z-statistics calculated for real and simulated artifacts, for spatial smoothing levels of the EPI time series varied between 0, 3 and 6 mm FWHM, as indicated on the horizontal axes. The time series smoothed with 6 mm were additionally analyzed using a version of the current injection scheme that was convolved with the Hemodynamic Response Function (HRF), replicating a standard approach for BOLD fMRI analyses. The linear fits were evaluated for different thresholding levels (0, 1.65, 2.3 and 3.1) of the simulated data (indicated by 0...3.1 on the upper horizontal axes). **A)** tDCS results for 1, 2 and 3.5 mA. In line with Figure S10, the lead configuration for real tDCS resulted in weak remote effects, so that correlations between simulated and measured effects remained lower than for the upper slices even at high current intensities. **B)** Loop results for 1 and 2 mA. The loop configuration caused stronger remote effects, which is apparent from the good correlations at 2 mA for all participants.

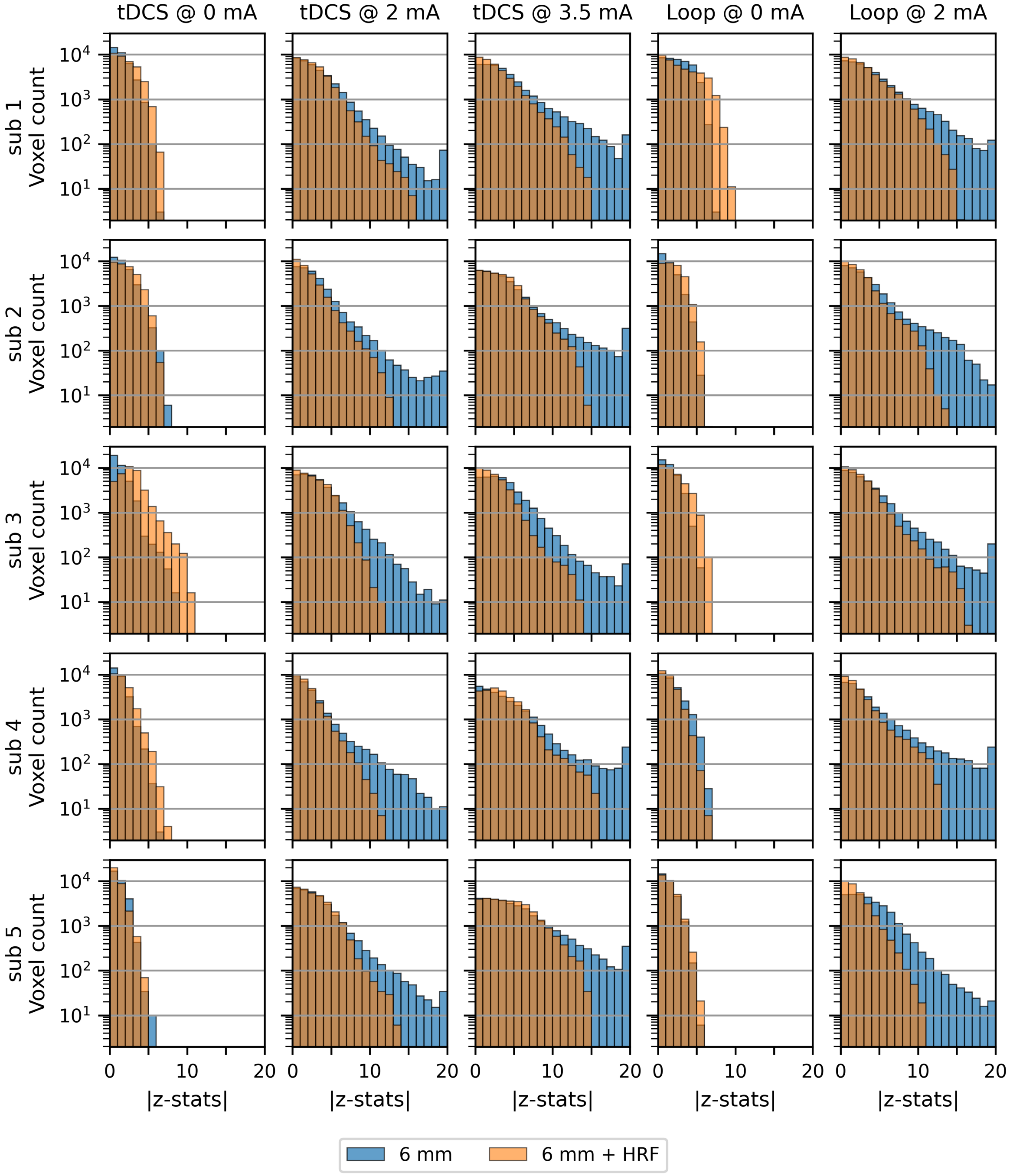


**Figure S13. Histograms of the voxel-wise activations determined for real fMRI data in the *upper* half of the FoV, for all subjects.** The horizontal axes depict the absolute values of the z-statistics from first level analyses of the current injection scheme, with the last histogram bin representing values $\geq$ 19. Results are shown for the highest current intensities applied with tDCS (2 and 3.5 mA) and loop (2 mA), where clear artifact-related "activations" were observed in all subjects. The 0 mA data is also included for comparison. Only the results for 6 mm (FWHM) smoothing are shown here, for analyses without (blue) and with (orange) convolution of the current injection scheme with the Hemodynamic Response Function (HRF). Longer tails (i.e., more voxels with high z-values) were observed for 0 and 3 mm smoothing in the acquisitions with applied currents (not shown).

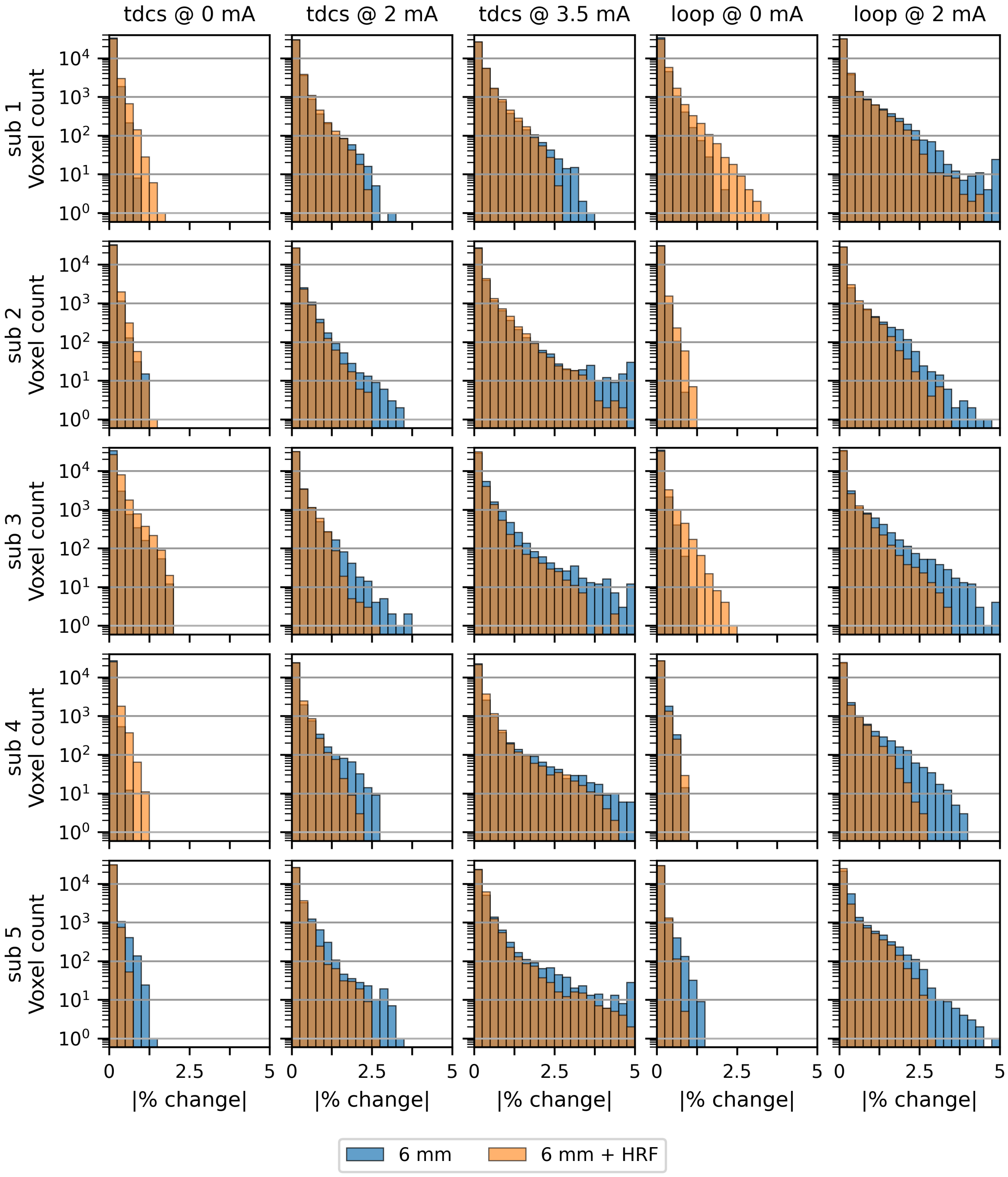


**Figure S14. Histograms of the voxel-wise percent signal changes determined for real fMRI data in the *upper* half of the FoV, for all subjects.** The last histogram bin includes all voxels with an absolute percent signal change $\geq 4.75$. Results are shown for the highest current intensities applied with tDCS (2 and 3.5 mA) and loop (2 mA), where clear artifact-related signal variations were observed in all subjects. The 0 mA data is also included for comparison. Only the results for 6 mm (FWHM) smoothing are shown here, for analyses without (blue) and with (orange) convolution of the current injection scheme with the Hemodynamic Response Function (HRF).

#### Supporting Material S8: Artifact simulation — group level analysis

Group level fMRI analyses were performed to verify the consistency of the current-induced signal changes across all five subjects. The EPI series were non-linearly co-registered to the MNI152 template using FSL FNIRT before conducting first-level analyses as described in Section 2.4 of the main text. The results obtained for each subject were then taken to a second level in FSL FEAT, where they were tested for fixed effects due to the small sample size. This analysis was repeated for each stimulation condition, for series with real and simulated artifacts. Linear Deming regression was performed on the group-level activation maps determined for real and simulated data (cf. Section 2.6 of the main text), after applying a voxel-level threshold of $z = 3.1$ to the simulated activations. The results obtained for 6 mm (FWHM) smoothing and convolution of the current injection scheme with the Hemodynamic Response Function (HRF) are presented here for their relevance to the fMRI community.

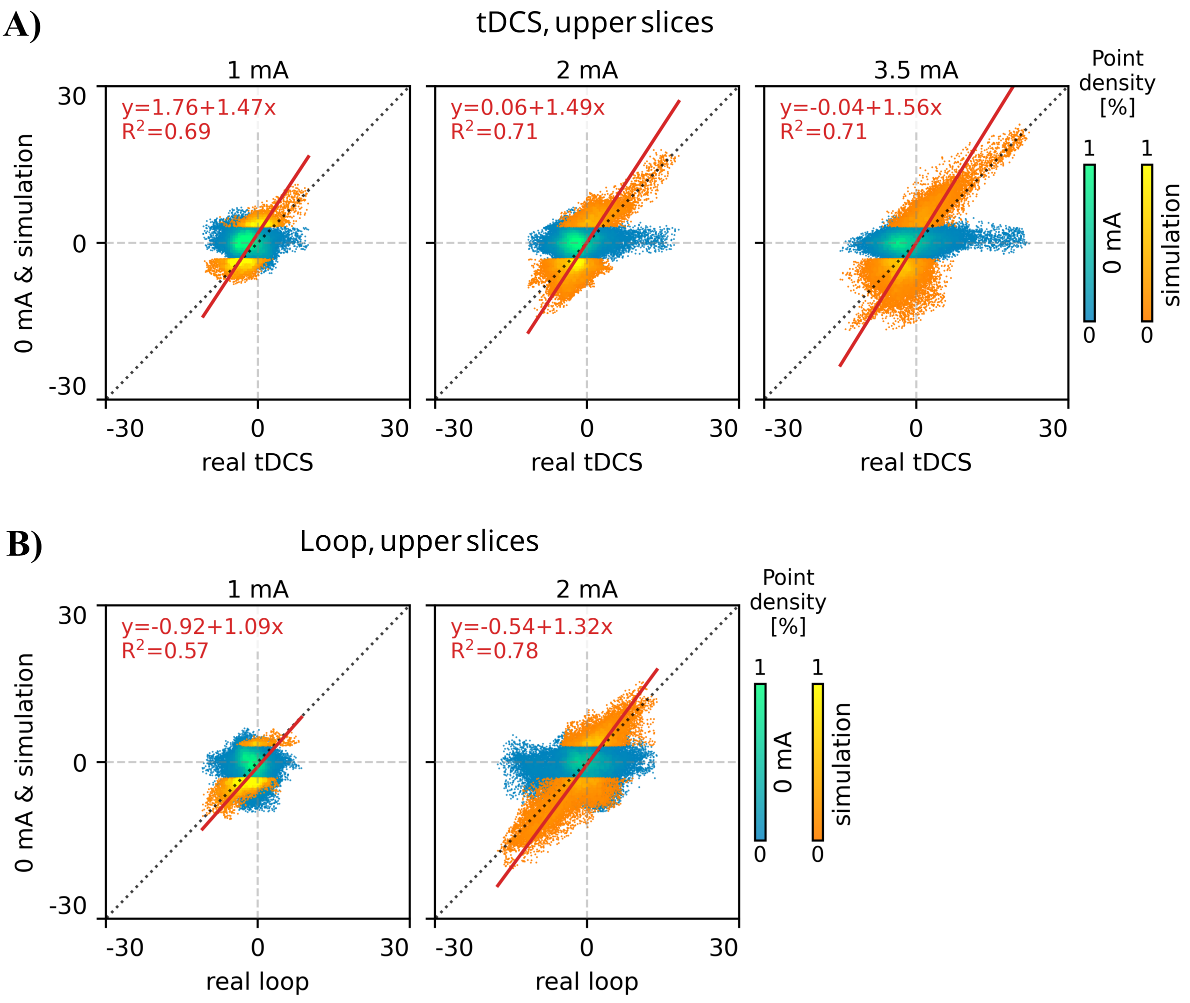


**Figure S15. Group-level (fixed effects) analysis of the current-induced signal changes in the *upper* half of the EPI FoV.** Shown are the scatter plots of the group-level activations for 0 mA and simulated tDCS/loop against real tDCS/loop, and linear fit between the simulated and real activations. The z-statistics of the simulated tDCS/loop were thresholded at 3.1 before computing the linear fit. **A)** tDCS results for 1, 2 and 3.5 mA. **B)** Loop results for 1 and 2 mA.

The good reproducibility of the artifact-related activations across subjects and the good accuracy of the simulations are quantitatively demonstrated in Figure S15. Significant linear relationships (with intercepts/slopes close to 0/1 and high $R^2$ values) between the activations determined for real and simulated artifacts were observed in all conditions, including the lowest current intensity (1 mA) in both tDCS and loop. Figure S16 reveals the underlying activation maps in one example slice, where the large similarity between real and simulated artifacts can be qualitatively assessed.

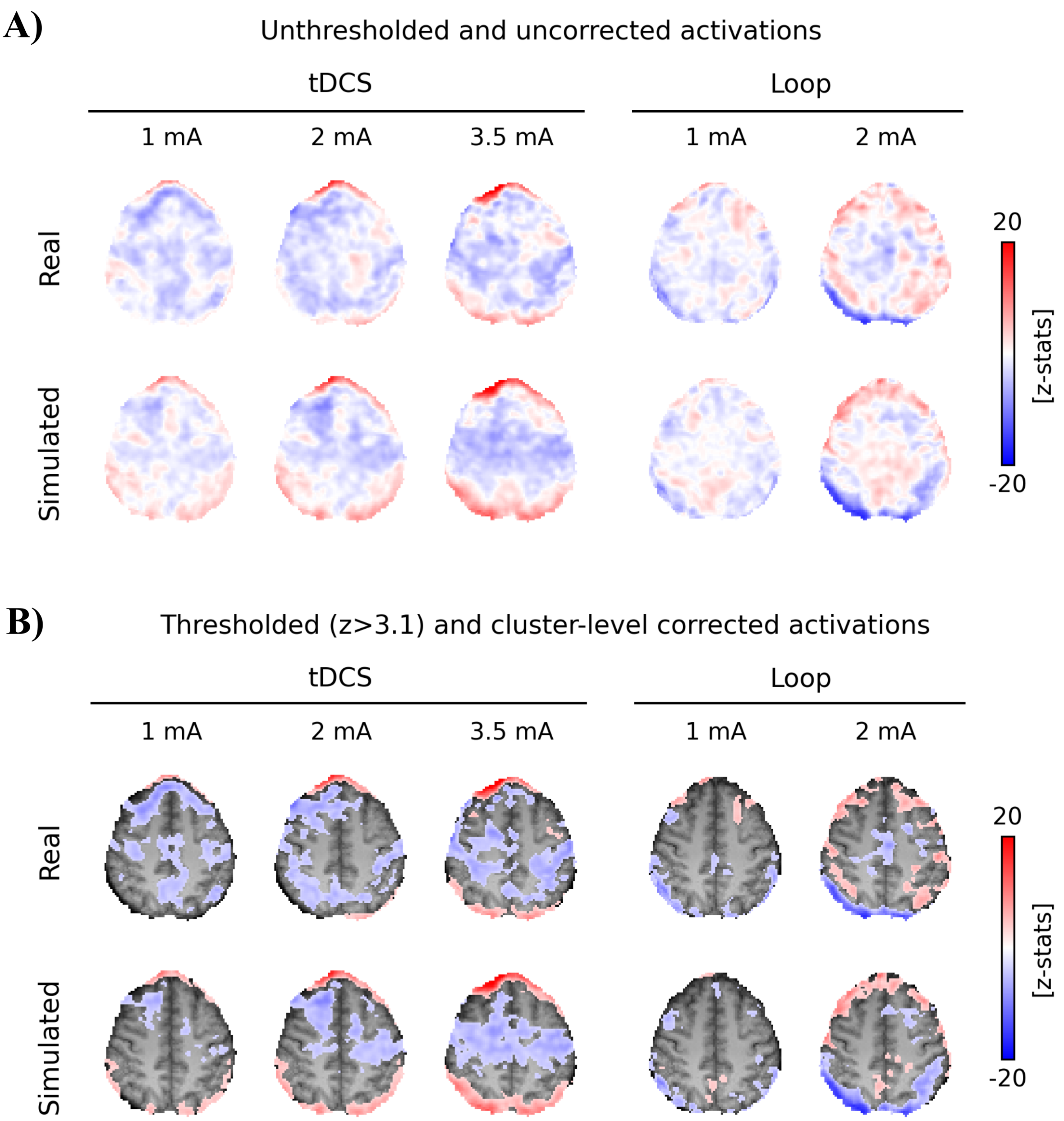


**Figure S16. Group-level (fixed effects) activations in an example slice.** Shown are the z-statistics maps computed for both real and simulated tDCS and loop at different current intensities, overlaid on the MNI template. **A)** Voxel-level z-statistics prior to thresholding. **B)** Activation clusters determined for a z-threshold of 3.1 and a cluster p-threshold of 0.01.

#### References


[1] P. Jezzard and R. S. Balaban. "Correction for geometric distortion in echo planar images from B0 field variations". In: *Magnetic Resonance in Medicine* 34.1 (July 1995), pp. 65–73. DOI: 10.1002/mrm.1910340111.

[2] R. W. Brown et al. *Magnetic Resonance Imaging: Physical Principles and Sequence Design*. Second Edition. Wiley Blackwell, 2014. Chap. 7, pp. 95–111. DOI: 10.1002/9781118633953.

[3] D. J. Larkman and R. G. Nunes. "Parallel magnetic resonance imaging". In: *Physics in Medicine & Biology* 52 (2007), pp. 15–55. DOI: 10.1088/0031-9155/52/7/R01.

[4] S. F. Cauley et al. "Interslice leakage artifact reduction technique for simultaneous multislice acquisitions". In: *Magnetic resonance in medicine* 72.1 (2014), pp. 93–102. DOI: 10.1002/mrm.24898.

[5] O. Heid. "Method for the phase correction of nuclear magnetic resonance signals". In: *US Patent 6,043,651* (2000).

[6] S. J. Inati, M. S. Hansen, and P. Kellman. "A Fast Optimal Method for Coil Sensitivity Estimation and Adaptive Coil Combination for Complex Images". In: *Proc. Intl. Soc. Mag. Reson. Med. 22*. 4407. 2014.

[7] D. O. Walsh, A. F. Gmitro, and M. W. Marcellin. "Adaptive Reconstruction of Phased Array MR Imagery". In: *Magnetic Resonance in Medicine* 43.5 (2000), pp. 682–690. DOI: 10.1002/(SICI)1522-2594(200005)43:5<682::AID-MRM10>3.0.CO;2-G.

[8] C. Göksu et al. "Human in-vivo brain magnetic resonance current density imaging (MRCDI)". In: *NeuroImage* 171 (May 2018), pp. 26–39. DOI: 10.1016/j.neuroimage.2017.12.075.

[9] C. Göksu et al. "Sensitivity and resolution improvement for in vivo magnetic resonance current-density imaging of the human brain". In: *Magnetic resonance in medicine* 86.6 (Dec. 2021), pp. 3131–3146. DOI: 10.1002/mrm.28944.

[10] F. Gregersen et al. "MR imaging of the magnetic fields induced by injected currents can guide improvements of individualized head volume conductor models". In: *Imaging Neuroscience* 2 (Dec. 2024), pp. 1–15. DOI: 10.1162/imag_a_00176.

[11] T. Cunha et al. "Increased brain coverage and efficiency when measuring current-induced magnetic fields by use of simultaneous multi-slice echo-planar MRI". In: *PLOS ONE* 21.1 (Jan. 2026), pp. 1–13. DOI: 10.1371/journal.pone.0341731.

[12] G. C. Scott et al. "Sensitivity of magnetic-resonance current-density imaging". In: *Journal of Magnetic Resonance (1969)* 97.2 (Apr. 1992), pp. 235–254. DOI: 10.1016/0022-2364(92)90310-4.

[13] A. Antal et al. "Transcranial direct current stimulation over the primary motor cortex during fMRI". In: *NeuroImage* 55.2 (Mar. 2011), pp. 590–596. DOI: 10.1016/j.neuroimage.2010.11.085.

[14] L. M. Li et al. "Brain state and polarity dependent modulation of brain networks by transcranial direct current stimulation". In: *Human brain mapping* 40.3 (Feb. 2019), pp. 904–915. DOI: 10.1002/HBM.24420.

[15] G. E. Hagberg et al. "The effect of physiological noise in phase functional magnetic resonance imaging: from blood oxygen level-dependent effects to direct detection of neuronal currents". In: *Magnetic Resonance Imaging* 26.7 (Sept. 2008), pp. 1026–1040. DOI: 10.1016/J.MRI.2008.01.010.

[16] J. Ashburner et al. *SPM12 Manual*. Ed. by The FIL Methods Group (and honorary members). Wellcome Centre for Human Neuroimaging, 2021.

[17] M. Jenkinson et al. “Improved Optimization for the Robust and Accurate Linear Registration and Motion Correction of Brain Images”. In: *NeuroImage* 17.2 (Oct. 2002), pp. 825–841. DOI: 10.1006/nimg.2002.1132.

[18] N. Todd et al. “Evaluation of 2D multiband EPI imaging for high-resolution, whole-brain, task-based fMRI studies at 3T: Sensitivity and slice leakage artifacts”. In: *NeuroImage* 124 (Jan. 2016), pp. 32–42. DOI: 10.1016/j.neuroimage.2015.08.056.
